# Inequalities in behavioural risk factor prevalence between five post-war generational cohorts of working age, England

**DOI:** 10.1101/2024.03.28.24305026

**Authors:** Madhavi Bajekal, Shaun Scholes

## Abstract

**Background and objectives:** Long-term trends in health risk factor prevalence, and in inequalities, are often summarised using age-standardised point prevalence to allow for age distribution changes over time and between groups. Policies to effectively promote health require the decomposition of social change into its age-period-cohort components. As a first step, we provide a descriptive age-cohort analysis to identify patterns of generational change in key behavioural risk factors between and within post-war cohorts of working age in England.

**Data:** Cross-sectional Health Survey for England data for participants aged 25-60 years was pooled between 1994-2019 (n=153,172) to construct five decennial cohorts (1940s, 1950s, 1960s, 1970s, 1980s). Socioeconomic status was classified by neighbourhood deprivation quintiles using the Index of Multiple Deprivation. Five behavioural risk factors were analysed: cigarette smoking (current-and heavy-smoking); alcohol consumption (frequency and heavy drinking); obesity; meeting recommended levels of physical activity and fruit and vegetables consumption.

**Methods:** Sex-specific analyses were conducted. Log-binomial regression models quantified the magnitude and direction of change in (i) prevalence ratios (PRs) between cohorts adjusting for age and deprivation and (ii) deprivation-specific PRs within- and between-cohorts to examine changes in absolute and relative inequalities between generations.

**Results:** In more recent cohorts, decreases in prevalence, independent of age and deprivation, were observed for current smoking, frequency of alcohol consumption and heavy drinking, resulting in decreasing absolute inequalities. However, obesity levels, particularly among females, reached their highest levels in the youngest 1980s cohort.

Relative inequalities in current smoking (most-versus least-deprived quintiles) peaked in the 1950s cohort (Males: PR 2.79 (95% CI: *2.57-3.04*); Females: PR 2.81; 95% CI*: 2.60-3.05)*), decreased in the 1960s cohort (M: PR 2.00 (95% CI: *1.70-2.34*); F: PR 2.58 (95% CI: *2.40-2.77*)), and remained stable thereafter. Inequalities in heavy smoking persisted over time among current smokers.

Higher obesity levels in the most-versus least-deprived quintiles were generally persistent across all five cohorts, albeit with some suggestion of widening inequalities in the younger-versus older-cohorts in females (1940s cohort: PR 1.55 (95% CI: *1.40-1.72*); 1960s cohort: PR 1.87 (95% CI: 1*.73-2.01*)). This pattern was also observed for mean body mass index (BMI).

For heavy drinking, relative inequality remained stable. Relative inequalities in fruit and vegetable consumption were lower in more recent cohorts. Physical activity levels were similar across cohorts, with little evidence of inequalities.

**Conclusion:** Our analysis of generational change reveals credible signals of behavioural risk factor changes in levels and in inequalities over successive post-war cohorts of working age.

## Introduction

There is a substantial body of work that demonstrates the persistence of socioeconomic inequalities in mortality and its associated behavioural risk factors in England.^1,2^ Typically, these studies rely on point estimates derived from social surveys or vital statistics data. Annual estimates are typically age-adjusted and stratified by one or more socioeconomic indicators of disadvantage to analyse changes in risk factor prevalence and trends in inequalities between different sub-groups over time.

However, in this study we use a different approach. We compare (i) differences in the prevalence of known health behavioural risk factors between post-war birth cohorts and examine (ii) changes in inequalities by constructing generational (or pseudo-, or synthetic-) cohorts using a long series of annual, repeated cross-sectional health surveys.^3^ Unlike longitudinal birth cohorts that prospectively track changes in individuals’ health, generational cohorts offer a unique perspective by capturing the broader social changes occurring over time between cohorts using repeated cross-sectional data. This method works to alert policy makers to probable cohort effects as long as the survey data are representative of the population, and its members are not experiencing significant mortality, which could lead to selective survival bias.

Research conducted using prospective longitudinal studies has demonstrated that the adoption of healthy lifestyle behaviours during young adulthood can effectively postpone the onset and lower the incidence of chronic diseases.^4.5^ However, such longitudinal studies tend to be resource-intensive and typically concentrate on a single cohort. Generational cohort studies based on annual, repeated cross-sectional data offer a valuable complement to longitudinal research by zeroing-in on the behavioural drivers of health and in estimating the magnitude of any changes in prevalence between earlier and later-born cohorts.

This approach has the potential to inform proactive public health strategies and policy making, contributing to gains in health expectancy of successive generations as they age.^6^ Furthermore, for pension actuaries, gaining insight into the direction of change in these pivotal determinants between successive post-war cohorts of working age provides a more solid basis for calibrating annuity models that project future life expectancies, ultimately facilitating more precise financial planning for retirement and pensions.

Using a generational cohort approach, the key research questions of the present study are:

1. Has the prevalence of key behavioural risk factors changed between cohorts, independently of age and deprivation?
2. Has the magnitude of risk factor inequalities changed between cohorts?

## Methods

### Data and participants

The Health Survey for England (HSE) is a series of repeated, annual cross-sectional health examination surveys designed to monitor health conditions and health-related behaviours in a nationally representative sample of the non-institutionalised population. A new sample each year is randomly selected by address. The sample size and the health topic focus varies each year with certain ‘core’ modules retained every year.^7^

HSE data by single-year of age are accessible through the End User Licence (EUL) via the UK Data Service (1994-2013). However, in the more recent EUL datasets (2014-2019), participants’ age is grouped, limiting its use for constructing a generational cohort series.

#### Birth cohorts and age

We aggregated annual cross-sectional HSE data from 1994 to 2019. This pooled data allowed us to categorise participants into five decennial post-war birth cohorts: the ’1940s’ cohort (born between 1940-49), the ’1950s’ cohort (born between 1950-59), the ’1960s’ cohort (born between 1960-69), the ’1970s’ cohort (born between 1970-79), and the ’1980s’ cohort (born between 1980-89).

We restricted analyses to participants aged 25 to 60 at the time of the survey. This age truncation resulted in varying age ranges for each cohort, spanning from 25 to 39 years in the youngest 1980s cohort to 45 to 60 years in the oldest 1940s cohort. The selected age-range of 25 to 60 years ensured that the analytical sample included participants of the same age in overlapping cohorts. The 1960s cohort covered nearly the full age range of the study, from 25 to 59 years (see Table 1). To put in context, in 2019 the central age of the five cohorts was 74, 64, 54, 44, and 34 years (plus/minus 5 years), from the oldest (1940s) cohort to the youngest (1980s). The total study sample size was 153,572. The mean age of the analytical sample was 41 years.

**Table 1:**
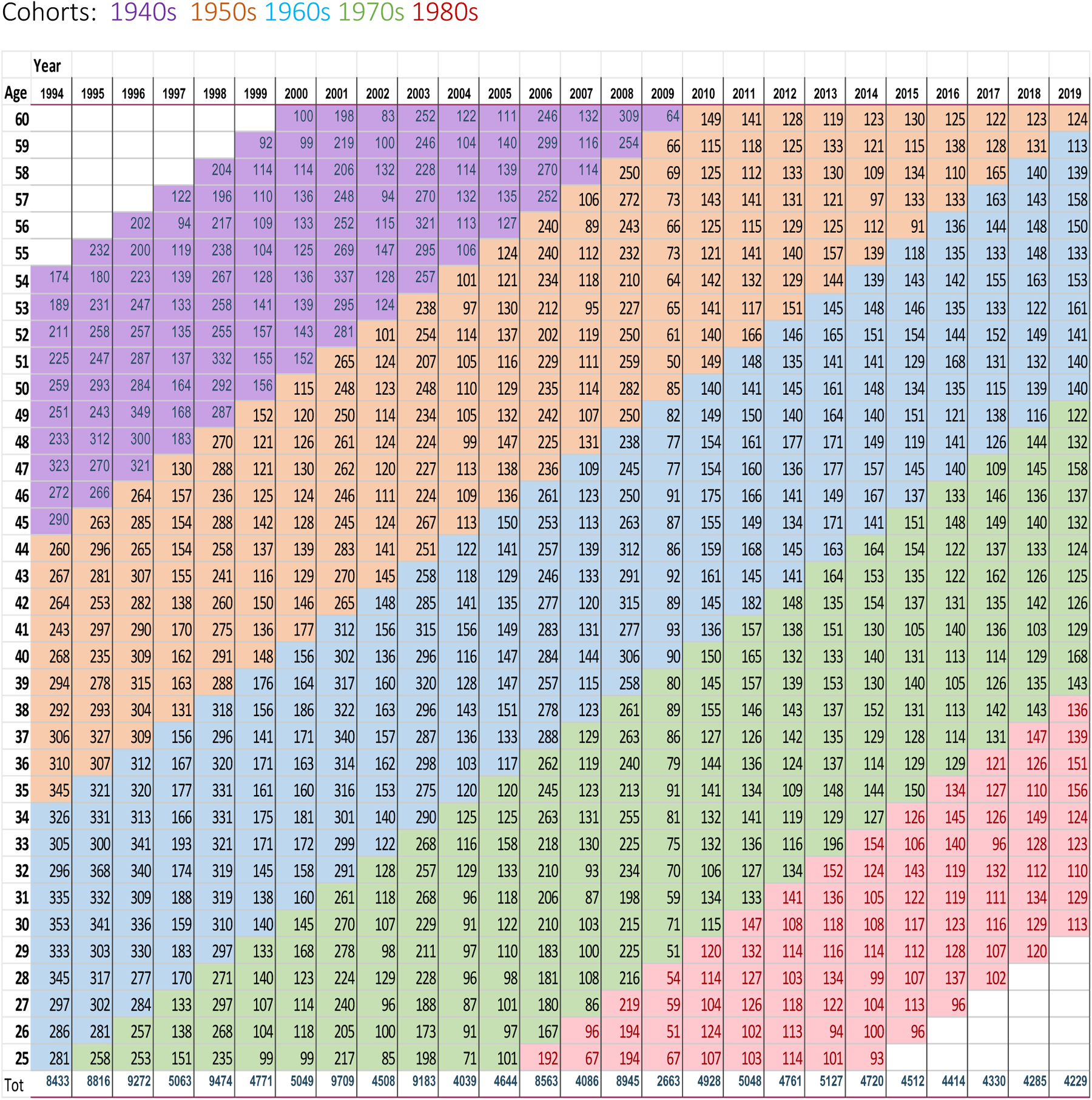
Lexis diagram, sample counts age by year (persons)

With the exception of the extreme tails truncated by the specified age cut-offs, we derived risk factor prevalence estimates by single-year of age by aggregating both the numerators and denominators over ten consecutive years of annual survey data for each cohort (see Supplementary Appendix S1_ Section B.1). The median sample size per single year of age for the 1960s cohort was 1,513.

Our decision to use 10-year grouping to construct the generational cohorts, rather than the more conventional 5-year grouping commonly used in demography, was mainly because of limits to sample sizes once the analytic sample is partitioned by age, sex and deprivation quintiles (see below) to assess inequalities in risk factor prevalence within-and between-cohorts.

### Behavioural risk factors

Over the last two decades, there have been changes in survey questions or updates to measurement methods to align with policy recommendations. Our primary aim has been to maintain a consistent definition of the chosen behavioural risk factors throughout the study years, thereby ensuring that we avoid introducing artificial ‘jumps’ in prevalence levels between cohorts. However, in cases where the variables were not collected consistently and could not be harmonised (e.g. changes to units of alcohol), or when data collection commenced more recently (e.g. fruit and vegetable consumption), we initiated the analysis from the first available year when these variables were consistently collected.

We selected five modifiable behavioural risk factors for our analysis, namely the proportions of individuals who:

- Were current cigarette smokers; and among them, those who were ‘heavy smokers’, defined as smoking 20 or more cigarettes per day.
- Typically consumed alcohol on five or more days per week over the past 12 months (frequency); and of those who consumed alcohol last week, those who were ‘heavy drinkers’ (8+ units for males and 6+ units for females) on the heaviest drinking day in the last week (amount).
- Had a measured Body Mass Index (BMI) of 30kg/m^2^ or more (i.e. obesity).
- Were physically active, defined as being engaged in 30 minutes or more of moderate- or vigorous-intensity activity on at least five days per week (including activity whilst at work).
- On average, consumed five or more portions of fruits and vegetables per day.

Further details of the survey protocol and the questions asked are provided in S1_Section A.

### Socio-economic circumstances

The HSE includes three measures of individual socio-economic position – namely, household income, highest educational attainment, and occupation-based social class (e.g. Registrar General’s social class, and the National Statistics Socio-Economic Classification, NSSEC). However, household income and educational attainment were collected on a consistent basis only from 1997 onwards. Additionally, there was a gap in the archiving of social class information in 2010-2011, resulting in a discontinuity in the series.

Aside from being incomplete series, the measurement of individual social position was also impacted by a substantial amount of item non-response, which affected cohorts differentially. For instance, information on household income was missing for approximately 30% of the analytic sample; but this ranged from 44% in the 1940s cohort to 16% in the 1980s cohort. Likewise, the percentage of missing data for the oldest and youngest cohorts was 23% and 0% for educational attainment, respectively. The equivalent figures were 2% and 18% respectively for social class.

In contrast, the extent of missing data for a geographical measure of relative deprivation based on the area of residence of the HSE participants, known as the Index of Multiple Deprivation (IMD) at the Lower Layer Super Output Area (LSOA),^8^ exhibited the lowest overall rate of missingness, standing at 7%. Notably, this missingness pattern did not vary significantly by cohort, ranging from 8% for the 1940s cohort to 2% for the 1980s cohort (See S1_SB.2).

Throughout the study’s duration, the national IMD scores have been periodically recalibrated to reflect changes in relative deprivation, with assessments conducted at various time points (e.g., IMD 2001, IMD 2007, IMD 2010, IMD 2015). Although certain small areas may have experienced shifts in their specific rankings over time, our approach has been to use the deprivation measure that was in effect at the time of each survey. Deprivation quintiles are sizeable and encompass roughly equal fifths of the national population. Importantly, the majority of LSOAs have consistently maintained their quintile-membership across time, indicating remarkable stability.^9^ In our analysis, we classify IMD1 as the most deprived quintile (Q1) and IMD5 as the least deprived quintile (Q5).

Our pragmatic selection of IMD quintiles as the primary measure of social disadvantage in this analysis is rooted in the theory that measures of area disadvantage encompass both compositional aspects (such as the concentration of deprived individuals) and contextual factors (such as the availability of employment opportunities) related to relative disadvantage.^10^

### Statistical Analyses

Sex-specific analyses were conducted. We estimated the prevalence of each behavioural risk factor by single year of age separately in each cohort, and present prevalence using three-year moving averages. Log-binomial regression models were used to estimate prevalence ratios (PR) using the specific binary risk factor as the dependent variable with age (continuous), cohort (categorical) and IMD quintile (categorical) as independent variables.^11^

Two sets of models were fitted. To examine cohort differences (research question 1), the first set of models (model 1) contained the main effects of cohort (1960s as reference), age and IMD (least deprived quintile as reference) to estimate PRs comparing the cohorts after adjustment for age and deprivation. To examine differences in the magnitude of relative inequalities between cohorts (research question 2), the second set of models (model 2) included an additional cohort by IMD interaction term (1940s cohort as reference). An overall test of the interaction term (16 terms) was used to jointly examine whether the PRs comparing the deprivation quintiles varied by cohort (after age adjustment). A more specific test of four terms was used to examine whether the relative gap in risk factor prevalence between the least and most deprived quintiles varied by cohort.

To facilitate interpretation of the patterns in inequalities, the second set of models was used to predict risk factor prevalence by deprivation quintiles within each cohort at age 40. We report on two measures of inequality – absolute and relative. Absolute inequality is the difference in prevalence between the most and least deprived quintiles (Q1-Q5); relative inequality is the ratio of the prevalence of the most deprived quintile to the least deprived (Q1/Q5). Examining changes in inequalities on both the absolute and relative scales is important. For example, if risk factor levels increase over time in the most- and least-deprived areas at about the same pace, relative inequalities between successively younger cohorts remain unchanged, but absolute inequalities could increase.

BMI was examined as a continuous measure. Linear regression models were used to estimate differences in mean BMI between cohorts after adjustment for age and deprivation (model 1) and quantify the magnitude of absolute inequalities within- and between-cohorts (model 2).

Data set preparation was performed in SPSS V.24.0 (IBM, Armonk, New York); analysis was performed in Stata V.17.1 (StataCorp LLC, College Station, Texas).

#### Weighting and missing data

In its early years the HSE was considered to be self-weighting, unless the sample sizes for a specific age or social group, such as children or minority ethnic groups, had been boosted in that year. Since 2003, non-response weights have been included. Notably, these non-response weights were introduced almost at the mid-point of the survey years between 1994 and 2019 analysed in this study. Consequently, nearly all data points for the 1940s cohort in our study are unweighted, while all data points for the 1980s cohort are weighted. The remaining cohorts represent a mix of weighted and unweighted data.

Given this inconsistency in cohort-specific weighting, we have opted not to incorporate survey non-response weights into this analysis (boost samples were not included and so the analytical sample is self-weighted). Previous studies that analysed generational cohorts using HSE data conducted sensitivity analyses with and without non-response weights, noting no significant differences in estimates.^12,13^

We opted for a complete case analysis in our regressions for several reasons. First, both the selected outcome and predictor variables (age, sex, IMD) exhibited reasonably low levels of item non-response, in part due to data being collected at the interview stage rather than at the nurse visit. Secondly, as behavioural variables are self-reported, reliable imputation using the limited set of sociodemographic information recorded consistently was not feasible. Therefore, a complete case analysis was considered the most suitable approach for our study.

## Results

Table 2 shows the overall prevalence for each risk factor by survey year for adults aged 25 to 60 in the pooled HSE study sample.

**Table 2.** Risk factor prevalence: averages by year.

| Year | Base | Current smokers | Heavy smokers | Usually drink on 5+ days/ per week | Heavy drinkers | Obese (BMI 30 kg/m<sup>2</sup> +) | Mean BMI | Physically active | 5+ Fruits and Vegetables daily |
| --- | --- | --- | --- | --- | --- | --- | --- | --- | --- |
|  | N | % | % | % | % | % | Kg/m<sup>2</sup> | % | % |
| 1994 | 8433 | 31.2 | 37.3 | 16.3 | - | 14.8 | 25.8 | - | - |
| 1995 | 8816 | 31.4 | 37.9 | 18.8 | - | 16.4 | 26.1 | - | - |
| 1996 | 9272 | 32.2 | 39.1 | 20.1 | - | 17.2 | 26.2 | - | - |
| 1997 | 5063 | 30.8 | 37.6 | 21.1 | - | 18.7 | 26.5 | 31.5 | - |
| 1998 | 9474 | 30.4 | 36.9 | 22.3 | - | 19.6 | 26.6 | 35.8 | - |
| 1999 | 4771 | 30.0 | 34.9 | 18.1 | - | 20.7 | 26.7 | - | - |
| 2000 | 5049 | 29.4 | 35.7 | 18.0 | - | 21.7 | 26.9 | - | - |
| 2001 | 9709 | 28.5 | 35.0 | 19.8 | - | 23.2 | 27.1 | - | 27.3 |
| 2002 | 4508 | 29.8 | 33.6 | 18.7 | - | 23.8 | 27.1 | - | 25.0 |
| 2003 | 9183 | 28.3 | 34.8 | 18.8 | - | 23.7 | 27.2 | 33.8 | 25.7 |
| 2004 | 4039 | 26.6 | 32.5 | 19.2 | - | 24.6 | 27.2 | 33.6 | 26.3 |
| 2005 | 4644 | 28.3 | 31.3 | 19.7 | - | 25.1 | 27.3 | - | 29.4 |
| 2006 | 8563 | 25.4 | 29.2 | 17.8 | - | 25.3 | 27.4 | 38.0 | 31.5 |
| 2007 | 4086 | 25.2 | 25.5 | 15.6 | 33.5 | 26.1 | 27.3 | - | 30.9 |
| 2008 | 8945 | 24.6 | 27.5 | 16.3 | 33.3 | 26.0 | 27.4 | 39.7 | 28.7 |
| 2009 | 2663 | 25.3 | 28.3 | 14.2 | 32.9 | 25.2 | 27.4 | - | 26.6 |
| 2010 | 4928 | 22.6 | 27.0 | 13.7 | 32.6 | 27.9 | 27.7 | - | 26.3 |
| 2011 | 5048 | 23.6 | 22.2 | 12.3 | 31.7 | 25.4 | 27.5 | - | 27.2 |
| 2012 | 4761 | 22.5 | 22.6 | 11.9 | 30.1 | 25.7 | 27.5 | 41.5 | - |
| 2013 | 5127 | 23.5 | 20.6 | 11.0 | 31.1 | 26.8 | 27.6 | - | 27.6 |
| 2014 | 4720 | 21.2 | - | 10.1 | 28.8 | 27.5 | 27.7 | - | - |
| 2015 | 4512 | 20.2 | - | 10.2 | 29.2 | 28.4 | 27.7 | - | - |
| 2016 | 4414 | 20.4 | - | 10.0 | 29.5 | 28.9 | 27.8 | - | - |
| 2017 | 4330 | 19.6 | - | 9.1 | 28.7 | 31.5 | 28.2 | - | - |
| 2018 | 4285 | 19.2 | - | 10.6 | 29.6 | 31.0 | 28.1 | - | - |
| 2019 | 4229 | 16.8 | - | 8.4 | 31.8 | 32.0 | 28.2 | - | - |
| Mean<sup>3</sup> | - | 26.5 | 33.0 | 16.2 | 31.2 | 23.4 | 27.1 | 36.4 | 27.8 |
| **N** | **153572** | **153290** | **35368<sup>1</sup>** | **150204** | **38914<sup>2</sup>** | **135854** | **135854** | **135682** | **49872** |

Notes: <sup>1</sup> Heavy smokers: selected base of all who were current smokers. <sup>2</sup> Heavy drinkers: selected base of all who had consumed alcohol in the last week. <sup>3</sup> Overall mean for pooled sample.

For each behavioural risk factor, we present a set of six figures, three for each sex displayed side-by-side to facilitate comparison.

An overview of these graphical representations is provided below:

### Percentage prevalence by cohort and age (top panel)

The first graph in the set shows the observed risk factor prevalence by cohort and single-year of age, smoothed using three-year moving averages. This helps to illustrate common age-patterns and, where ages overlap between successive cohorts, the between-cohort differences.

### Cohort-specific prevalence ratios with 1960 as reference cohort (middle panel)

In this, based on the first set of log-binomial models, we present the estimated PRs comparing the cohorts after adjustment for age and deprivation, with associated 95% confidence intervals (95% CIs). The 1960s cohort was chosen as reference. To facilitate comparison between cohorts, the values on the bar graph have been rebased with the 1960 reference value of 1 set to zero. Hence an estimated PR of 0.20 for a specific cohort indicates that the prevalence was estimated to be 20% higher (in relative terms) than for the 1960s cohort independently of age and deprivation.

### Predicted prevalence at age 40 by cohort and deprivation quintile (bottom panel)

The third graph in the set, based on the second set of log-binomial models (containing a cohort by IMD interaction term), displays model-predicted risk factor prevalence at age 40, by IMD quintile and cohort. Additionally, the second y-axis displays the estimated PR between the most and least deprived quintiles within each cohort (least deprived as reference). Hence, both the difference in prevalence between the most and least deprived quintiles in each cohort (*absolute inequality; Q1-Q5*) and the ratio between them (*relative inequality; Q1/Q5*) are presented.

Tables that support the data presented in each figure can be found in the Supplementary Appendix, (S1_ SC and SD). A summary table of the estimated PRs (most versus least deprived quintiles) for each binary risk factor at age 40, by sex and cohort is presented in Table 3. Results for mean BMI are presented separately in S1_SD6.

**Table 3:** Relative Inequality in prevalence ratios (most versus least deprived quintiles) in behavioural risk factors at age 40, by sex and post-war birth cohorts (with 95% CIs).

|  | Cohort |  |  |  |  |
| --- | --- | --- | --- | --- | --- |
|  | 1940s | 1950s | 1960s | 1970s | 1980s |
| **Current smokers** |  |  |  |  |  |
| Males | 2.62 | 2.79 | 2.19 | 1.95 | 2.00 |
| 95% CI | 2.34,2.92 | 2.57,3.04 | 2.04,2.35 | 1.78,2.14 | 1.70,2.34 |
| Females | 2.35 | 2.81 | 2.58 | 2.38 | 2.27 |
| 95% CI | 2.12,2.60 | 2.60,3.05 | 2.40,2.77 | 2.16,2.63 | 1.92,2.68 |
| **Heavy smoking (20+ per day)** |  |  |  |  |  |
| Males | 1.23 | 1.26 | 1.37 | 1.31 | 1.34 |
| 95% CI | 1.08,1.39 | 1.12,1.41 | 1.21,1.56 | 1.07,1.61 | 0.71,2.54 |
| Females | 1.34 | 1.72 | 2.01 | 2.16 | 2.55 |
| 95% CI | 1.15,1.57 | 1.48,1.99 | 1.70,2.38 | 1.62,2.90 | 1.05,6.20 |
| **Drink alcohol on 5+ days per week** |  |  |  |  |  |
| Males | 0.75 | 0.68 | 0.72 | 0.70 | 0.89 |
| 95% CI | 0.68,0.83 | 0.63,0.75 | 0.65,0.78 | 0.61,0.81 | 0.67,1.20 |
| Females | 0.49 | 0.53 | 0.55 | 0.56 | 0.57 |
| 95% CI | 0.42,0.57 | 0.47,0.60 | 0.49,0.62 | 0.46,0.67 | 0.39,0.84 |
| **Heavy drinking (units drank on heaviest drinking day in the last week)** |  |  |  |  |  |
| Males (8+ units) | 1.10 | 1.31 | 1.24 | 1.16 | 1.03 |
| 95% CI | 0.69,1.75 | 1.13,1.52 | 1.11,1.39 | 1.03,1.31 | 0.90,1.19 |
| Females (6+ units) | 1.11 | 1.34 | 1.42 | 1.54 | 1.30 |
| 95% CI | 0.59,2.10 | 1.10,1.62 | 1.25,1.61 | 1.35,1.75 | 1.13,1.50 |
| **Obesity (BMI 30+ kg/m<sup>2</sup>)** |  |  |  |  |  |
| Males | 1.29 | 1.24 | 1.17 | 1.17 | 1.13 |
| 95% CI | 1.15,1.45 | 1.14,1.35 | 1.08,1.27 | 1.04,1.30 | 0.94,1.36 |
| Females | 1.55 | 1.75 | 1.87 | 1.83 | 1.85 |
| 95% CI | 1.40,1.72 | 1.62,1.90 | 1.73,2.01 | 1.66,2.03 | 1.58,2.16 |
| **Physically active (30+ mins moderate or vigorous, 5+ days per week)** |  |  |  |  |  |
| Males | 0.92 | 1.01 | 1.03 | 1.12 | 1.02 |
| 95% CI | 0.79,1.08 | 0.90,1.14 | 0.94,1.13 | 1.00,1.25 | 0.84,1.26 |
| Females | 0.89 | 0.94 | 0.88 | 1.05 | 0.90 |
| 95% CI | 0.74,1.06 | 0.84,1.06 | 0.80,0.98 | 0.92,1.19 | 0.73,1.11 |
| **F&V 5+ portions per day** |  |  |  |  |  |
| Males | 0.52 | 0.67 | 0.81 | 0.74 | 0.69 |
| 95% CI | 0.43,0.63 | 0.59,0.77 | 0.71,0.91 | 0.65,0.85 | 0.52,0.92 |
| Females | 0.56 | 0.56 | 0.67 | 0.69 | 0.78 |
| 95% CI | 0.48,0.65 | 0.50,0.63 | 0.60,0.74 | 0.62,0.76 | 0.63,0.97 |

Notes: Ratios over 1 indicate prevalence in most deprived areas was greater than in the least deprived areas; ratios under 1 (eg Alcohol 5+ days pw) indicate prevalence lower in the most deprived vs affluent areas. See S1\_Tables SD1-SD8 for predicted prevalence ratios by IMD quintiles and model p values for interactions.

These visualisations and tables together offer an overview of the comparative differences in prevalence and in inequalities between cohorts associated with each risk factor, separately for males and females.

### Current Smoking

#### Age patterns and cohort differences

The age pattern of current smoking was consistent across cohorts. Prevalence was highest at age 25, and fell with increasing age for each cohort for both sexes (Fig 1, top panel). Proportions currently smoking were slightly lower in females than in males (S1_Table SC1).

**Fig 1.**
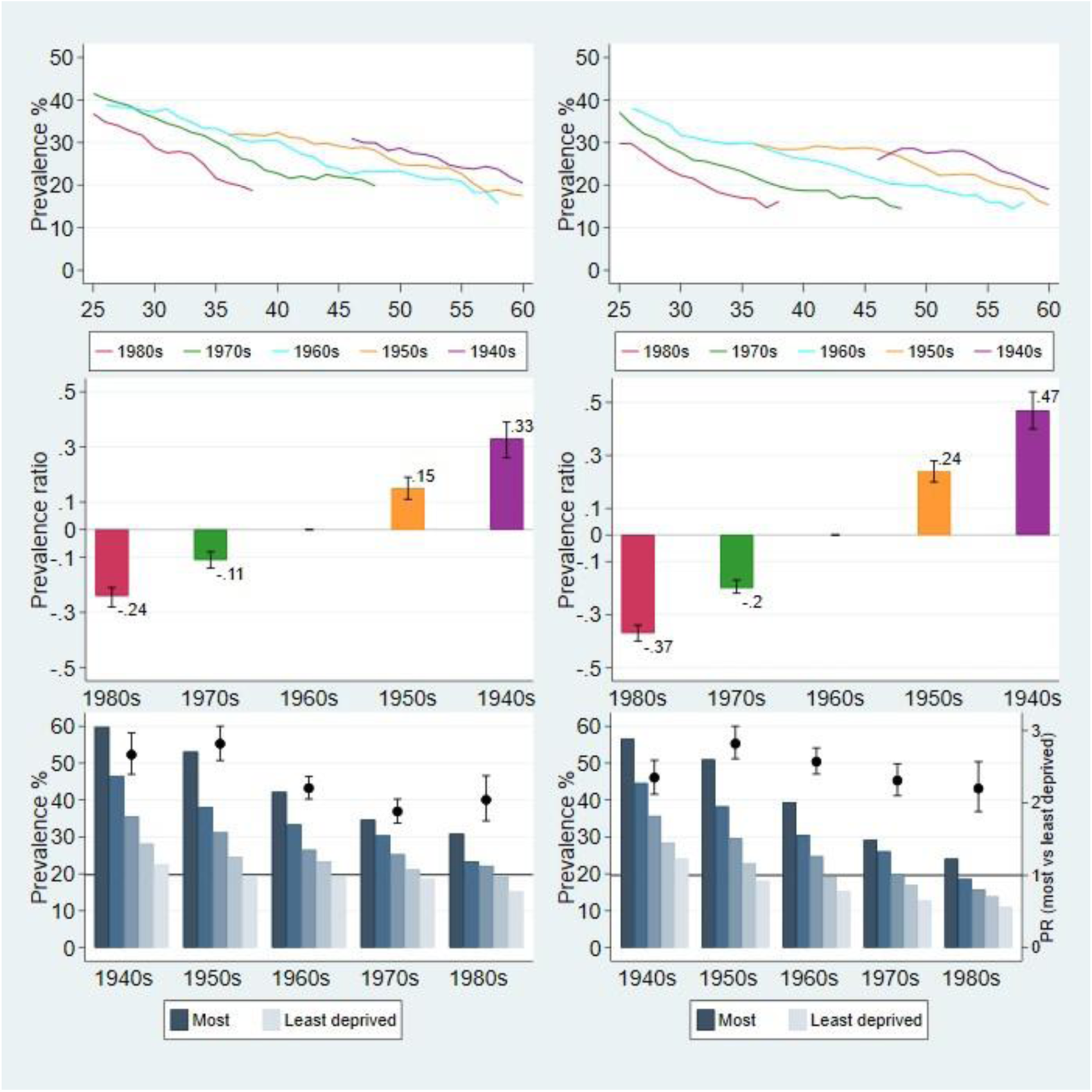
Current smoking. Notes: Males left panel, Females right panel. **Top panel**: 3-year moving averages of observed prevalence (%) by age and cohort. **Middle panel**: Prevalence ratio (value at top of bar), relative to the 1960s cohort (rebased to equal 0), with 95% confidence intervals (CIs) **Bottom panel**: Predicted prevalence (%) at age 40, by birth cohort and deprivation quintiles (primary y-axis); prevalence ratio (PR) between most and least deprived quintiles with 95% CIs (secondary y-axis). Unlike top and middle panels, cohorts are plotted from oldest (1940s) to youngest (1980s). PR greater than 1 indicates prevalence is higher in most versus least deprived quintiles; less than indicates prevalence is lower in most versus least deprived quintiles. PR is statistically significant at the 95% level when the CIs do not span 1.

As apparent from the stacking by age for overlapping cohorts, proportions of current smoking fell between successive cohorts, being lowest in the 1980s cohort and highest in the 1940s cohort (Fig 1). After adjusting for age and IMD composition of each cohort, relative to the 1960s cohort, prevalence rates for males were 24% lower in the 1980s cohort, and 33% higher in the 1940s cohort. For females, the difference was greater, at 37% lower and 47% higher, respectively.

#### Inequalities

Smoking prevalence was consistently higher in the most deprived compared to the least deprived quintiles across all cohorts. From its highest levels in the 1940s cohort, prevalence fell for successively younger cohorts in all deprivation quintiles and both sexes (Fig 1). Predicted prevalence at age 40 for each cohort halved for both males and females in the most deprived quintile between the oldest (1940s) and youngest (1980s) cohorts (from 60% to 31% for males; and 57% to 24% for females, see Table SD1). The percentage point fall between the 1940s and 1980s cohorts in the least deprived quintile was smaller, albeit from a lower base prevalence (males: 23% to 15%; females: 24% to 11%, respectively).

As a result, absolute inequality, as measured by the ‘gap’ or difference in smoking prevalence between the most and least deprived IMD quintiles in each successively younger cohort decreased. Relative inequality, as measured by the PR between the most and least deprived quintiles (least deprived as reference), was at its highest point in the 1950s cohort, at 2.80 (95% CI: 2.57-3.04) times higher for males and 2.82 (95% CI: 2.61-3.06) times higher for females (Fig 1, Table 2). This relative gap fell in the 1960s cohort and remained relatively stable thereafter. This suggests that the pace of fall in current smoking for successively younger cohorts was faster in the most deprived quintile compared to the least deprived quintile.

### Heavy smoking

Moreover, of those who were current smokers in each cohort, the prevalence of heavy smoking (20 or more cigarettes per day) also fell dramatically across all deprivation quintiles (see Fig 2, S1_ Tables SC2 and SD2). The PRs indicating the magnitude of relative inequality (above 1, indicating higher levels in the most versus least deprived quintiles) remained stable across successive cohorts, suggesting that the pace of fall in heavy smoking was of a similar magnitude across deprivation quintiles.

**Fig 2.**
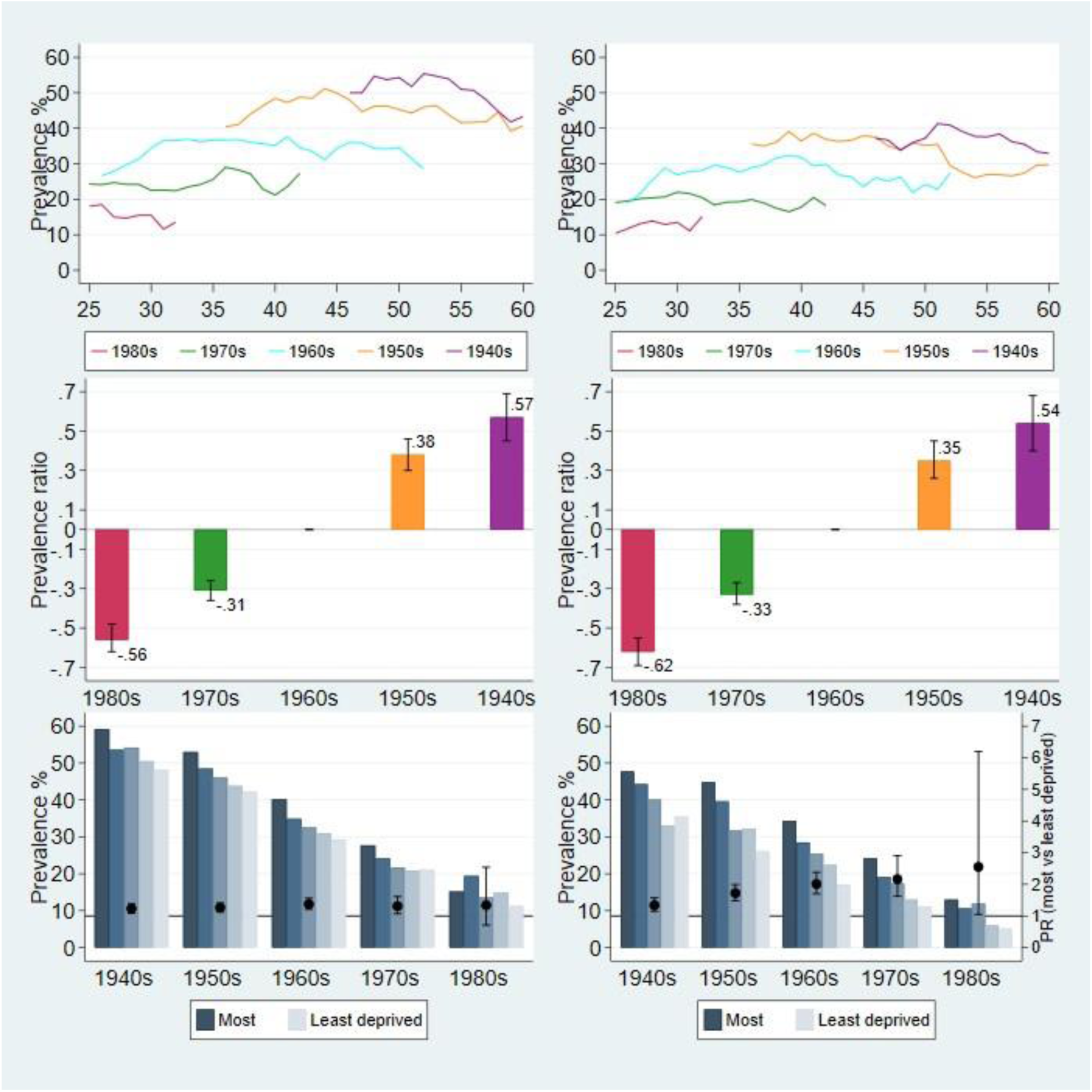
Heavy smoking (current smokers who smoked 20 or more cigarettes per day) Notes: Males left panel, Females right panel. **Top panel**: 3-year moving averages of observed prevalence (%) by age and cohort. **Middle panel**: Prevalence ratio (value at top of bar), relative to the 1960s cohort (rebased to equal 0), with 95% confidence intervals (CIs) **Bottom panel**: Predicted prevalence (%) at age 40, by birth cohort and deprivation quintiles (primary y-axis); prevalence ratio (PR) between most and least deprived quintiles with 95% CIs (secondary y-axis). Unlike top and middle panel, cohorts are plotted from oldest (1940s) to youngest (1980s). PR greater than 1 indicates prevalence is higher in most versus least deprived quintiles; less than 1 indicates prevalence is lower in most versus least deprived quintiles. PR is statistically significant at the 95% level when the CIs do not span 1.

### Alcohol consumption: frequency

#### Age patterns and cohort differences

The prevalence of frequent drinking (daily or up to five days a week) showed a marked decrease between successively younger cohorts for both sexes, particularly for males (Fig. 3, S1_Tables SC3).

Relative to the 1960s cohort, levels of frequent drinking were more than 50% higher for the 1940s cohort and more than 50% lower for the 1980s cohort.

**Fig 3.**
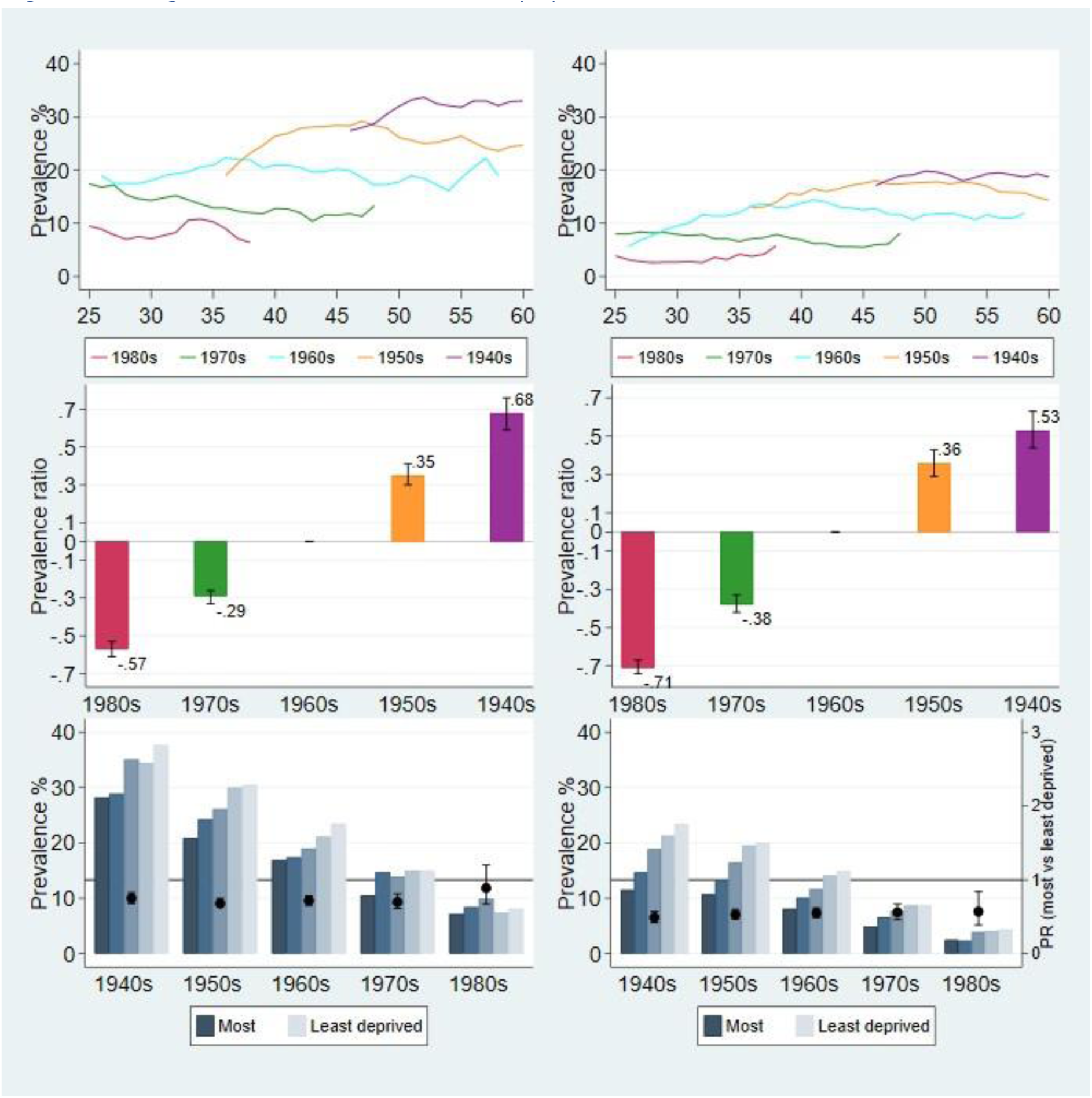
Drinking alcohol on five or more days per week. Notes: Males left panel, Females right panel. **Top panel**: 3-year moving averages of observed prevalence (%) by age and cohort. **Middle panel**: Prevalence ratio (value at top of bar), relative to the 1960s cohort (rebased to equal 0), with 95% confidence intervals (CIs) **Bottom panel**: Predicted prevalence (%) at age 40, by birth cohort and deprivation quintiles (primary y-axis); prevalence ratio (PR) between most and least deprived quintiles with 95% CIs (secondary y-axis). Unlike top and middle panel, cohorts are plotted from oldest (1940s) to youngest (1980s). PR greater than 1 indicates prevalence is higher in most versus least deprived quintiles; less than 1 indicates prevalence is lower in most versus least deprived quintiles. PR is statistically significant at the 95% level when the CIs do not span 1.

#### Inequalities

The predicted prevalence of frequent drinking at age 40 was consistently higher in the least deprived quintiles when compared to the most deprived (Fig 3). Females in the most deprived quintile were about half as likely, and males about 30% less likely, as those in the least deprived quintile to be frequent drinkers. Whilst the absolute gap between the most and least deprived quintiles declined between successively younger cohorts for both sexes, the relative gap remained stable across the cohorts (Fig 3, Table 2 and S1_Table SD3).

### Alcohol consumption: heavy drinking

#### Age patterns and cohort differences

The prevalence of heavy drinking (8+ units for males and 6+ units for females on the heaviest drinking day in the last week) among those who reported that they had consumed alcohol in the last week showed mixed results partly due to inconsistent data collection on units consumed in the first half of the survey period, which limited the age range for valid data for the older cohorts (Table 1). In comparison to the 1960s cohort, heavy drinking levels were notably lower for the 1970s and 1980s cohorts, for both sexes (see Fig 4), but were similar for the two oldest cohorts (1940s and 1950s).

**Fig 4.**
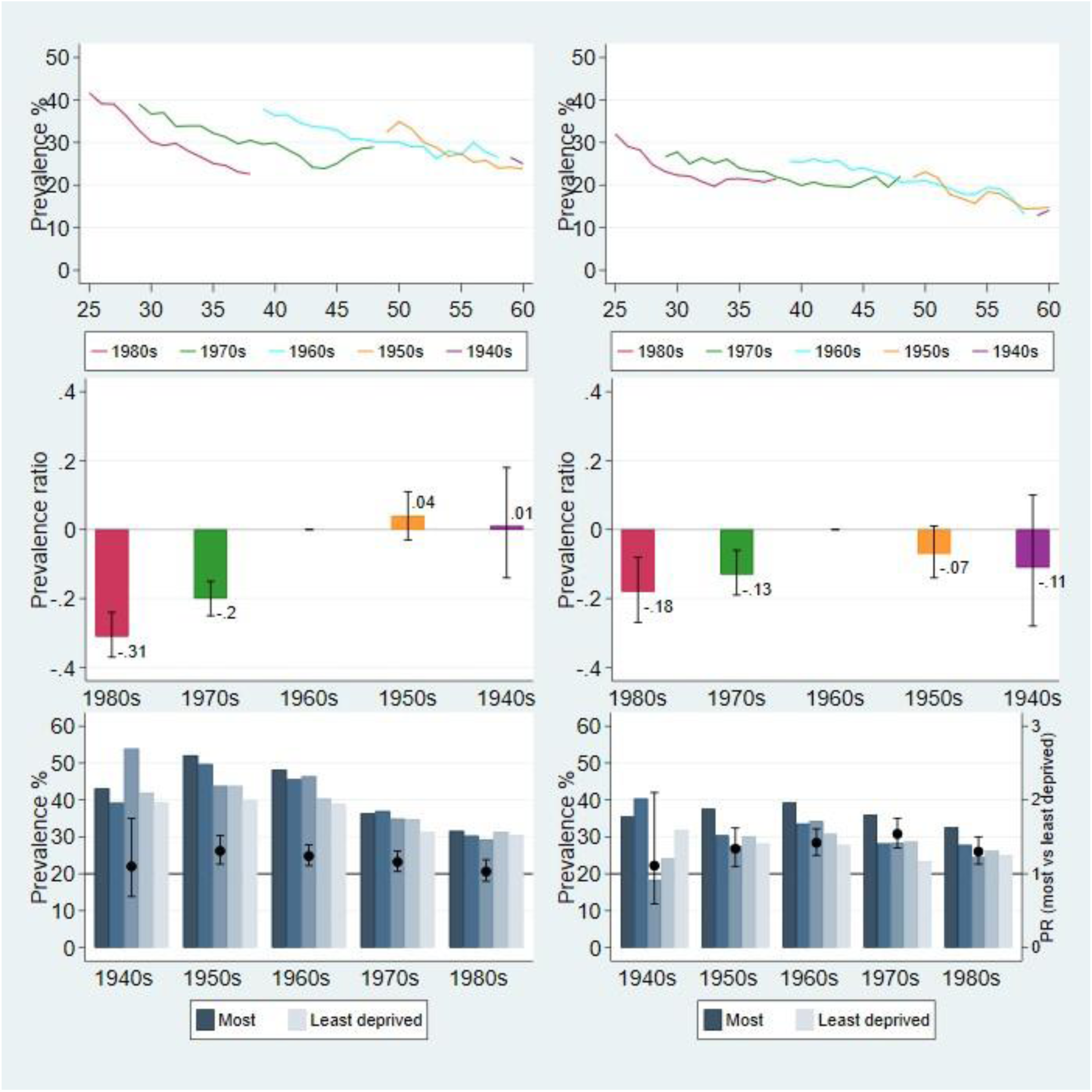
Heavy drinking: Drinkers consuming 8 units or more (males) or 6 units or more (females) on heaviest drinking day in the last week. Notes: Males left panel, Females right panel. **Top panel**: 3-year moving averages of observed prevalence (%) by age and cohort. **Middle panel**: Prevalence ratio (value at top of bar), relative to the 1960s cohort (rebased to equal 0), with 95% confidence intervals (CIs) **Bottom panel**: Predicted prevalence (%) at age 40, by birth cohort and deprivation quintiles (primary y-axis); prevalence ratio (PR) between most and least deprived quintiles with 95% CIs (secondary y-axis). Unlike top and middle panel, cohorts are plotted from oldest (1940s) to youngest (1980s). PR greater than 1 indicates prevalence is higher in most versus least deprived areas; less than 1 indicates prevalence is lower in most versus least deprived areas. PR is statistically significant at the 95% level when the CIs do not span 1.

#### Inequalities

In contrast to the pattern of frequent drinking, relative inequalities were evident for heavy drinking among all cohorts in both sexes, with higher levels in the most versus least deprived quintiles (Table 2; S1_Table SD4).

Whether analysed either from the perspective of consumption frequency or the amount consumed on the heaviest drinking day, it is evident that the 1980s cohort in particular showed the lowest levels of alcohol consumption across all deprivation quintiles.

### Obesity

#### Age patterns and cohort differences

The prevalence of obesity (BMI 30+ kg/m^2^) showed a steady increase across cohorts, with the lowest rates observed in the 1940s cohort and the highest in the 1980s cohort. This trend was especially pronounced among females (Fig 5). As expected, obesity levels increased consistently with advancing age, and these levels did not significantly differ between the sexes. After adjustment for age and deprivation, compared to the 1960s cohort, obesity levels in relative terms were 28% higher for males in the 1980s cohort and 37% lower for those in the 1940s cohort. For females, the corresponding levels were 41% higher for the 1980s cohort and 28% lower for the 1940s cohort.

**Fig 5.**
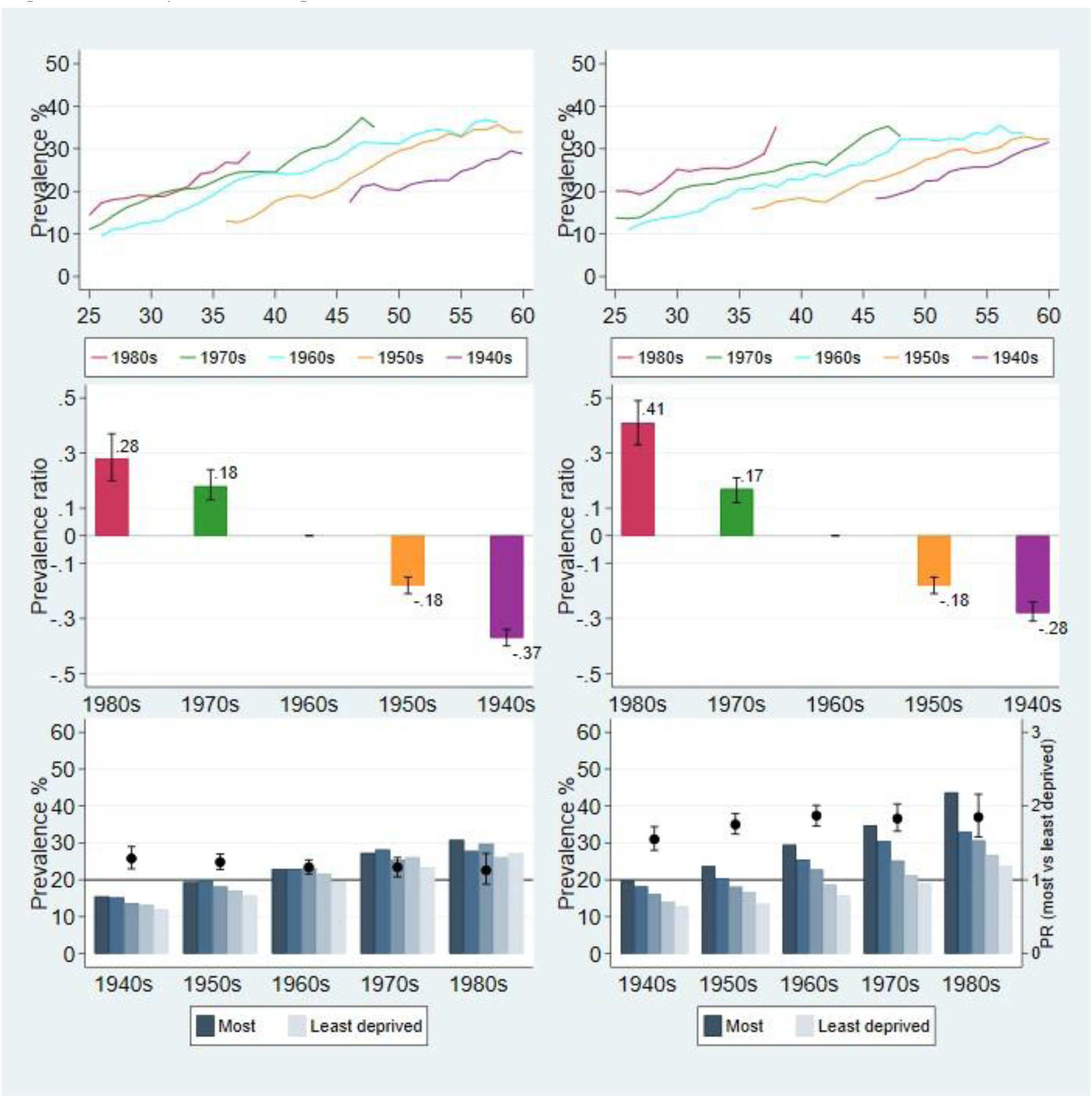
Obesity: BMI 30kg/m^2^ or above. Notes: Males left panel, Females right panel. **Top panel**: 3-year moving averages of observed prevalence (%) by age and cohort. **Middle panel**: Prevalence ratio (value at top of bar), relative to the 1960s cohort (rebased to equal 0), with 95% confidence intervals (CIs) **Bottom panel**: Predicted prevalence (%) at age 40, by birth cohort and deprivation quintiles (primary y-axis); prevalence ratio (PR) between most and least deprived quintiles with 95% CIs (secondary y-axis). Unlike top and middle panel, cohorts are plotted from oldest (1940s) to youngest (1980s). PR greater than 1 indicates prevalence is higher in most versus least deprived areas; less than 1 indicates prevalence is lower in most versus least deprived areas. PR is statistically significant at the 95% level when the CIs do not span 1.

#### Inequalities

The magnitude of absolute inequality in obesity within cohorts for males showed a 3-4 percentage point (pp) difference between the most and least deprived quintiles. For females, absolute inequality was not only higher but also increased between successively younger cohorts from 7 pp in the 1940s cohort to 20 pp in the 1980s cohort.

Predicted prevalence at age 40 consistently showed higher levels of obesity in the most deprived quintile for both sexes across all cohorts, except for males in the 1980s cohort, where the estimated PR did not attain statistical significance (PR 1.13; 95% CI: 0.94-1.36). The relative gap between the most and least deprived quintiles consistently and significantly exceeded 1 for females across all cohorts, with the PR increasing between the 1940s (PR 1.55; 95% CI: 1.40-1.72) to the 1960s cohort (PR 1.87; 95% CI: 1.73-2.01), and then remaining relatively stable thereafter (Table 2; S1_Table SD5).

### Mean BMI

#### Cohort differences

Rather than focusing solely on obesity at the upper end of the weight-to-height spectrum, an analysis of mean BMI confirmed the observed increase in higher BMI in the younger versus older cohorts (Fig 6).

**Fig 6.**
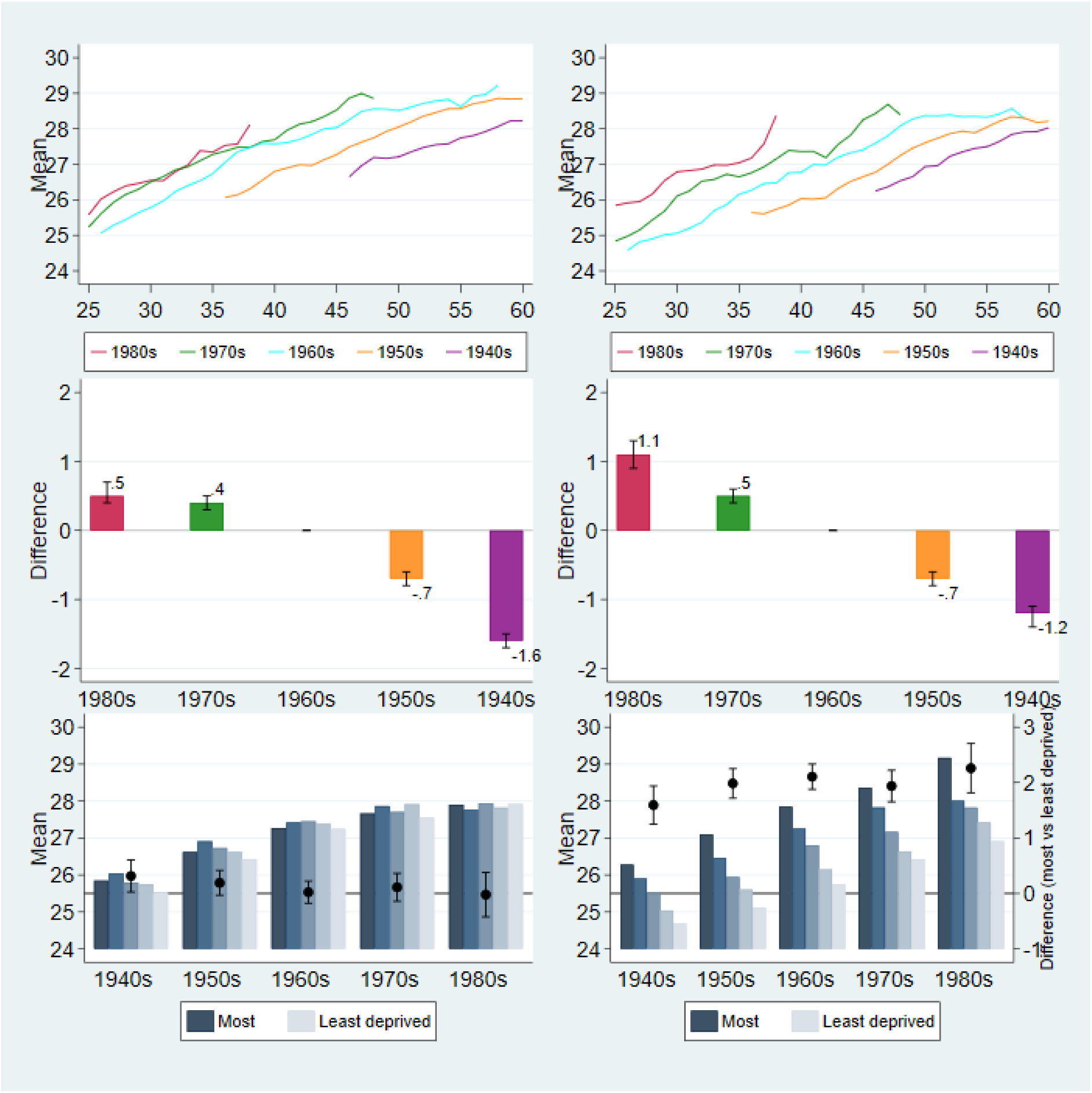
Mean BMI. Notes: Males left panel, Females right panel. **Top panel**: 3-year moving averages of mean BMI by age and cohort. **Middle panel**: Absolute difference in mean BMI (value at top of bar) relative to the 1960s cohort, with 95% confidence intervals (CIs) **Bottom panel**: Predicted mean BMI at age 40, by birth cohort and deprivation quintiles (primary y-axis); absolute difference in mean BMI between most and least deprived quintiles with 95% CIs (secondary y-axis). Unlike top and middle panel, cohorts are plotted from oldest (1940s) to youngest (1980s).

#### Inequalities

In males, higher levels of BMI in the most versus least deprived quintile were significantly higher only in the oldest (1940s) cohort (difference in mean BMI: 0.31kg/m^2^; 95% CI: 0.03-0.60) (S1_Table SD6). In contrast, in females, those in the most versus least deprived quintile had significantly higher mean BMI in each cohort, with the absolute gap increasing from 1.59kg/m^2^ (95% CI: 1.25-1.94) in the 1940s cohort to 2.26kg/m^2^ (95% CI: 1.82-2.71) in the 1980s cohort.

### Physical Activity

#### Age and cohort differences

At all ages between 25-60 years the proportion of physically active males (engaged in 30+ minutes of at least moderate-intensity physical activity, 5+ days/week) exceeded that of females, with levels highest at age 25 and declining gradually with advancing age. Age and deprivation-adjusted levels of sufficient physical activity were notably higher for participants in the youngest versus older cohorts, for both sexes (Fig 7). The difference between the youngest and oldest cohorts was more pronounced in females than in males.

**Fig 7.**
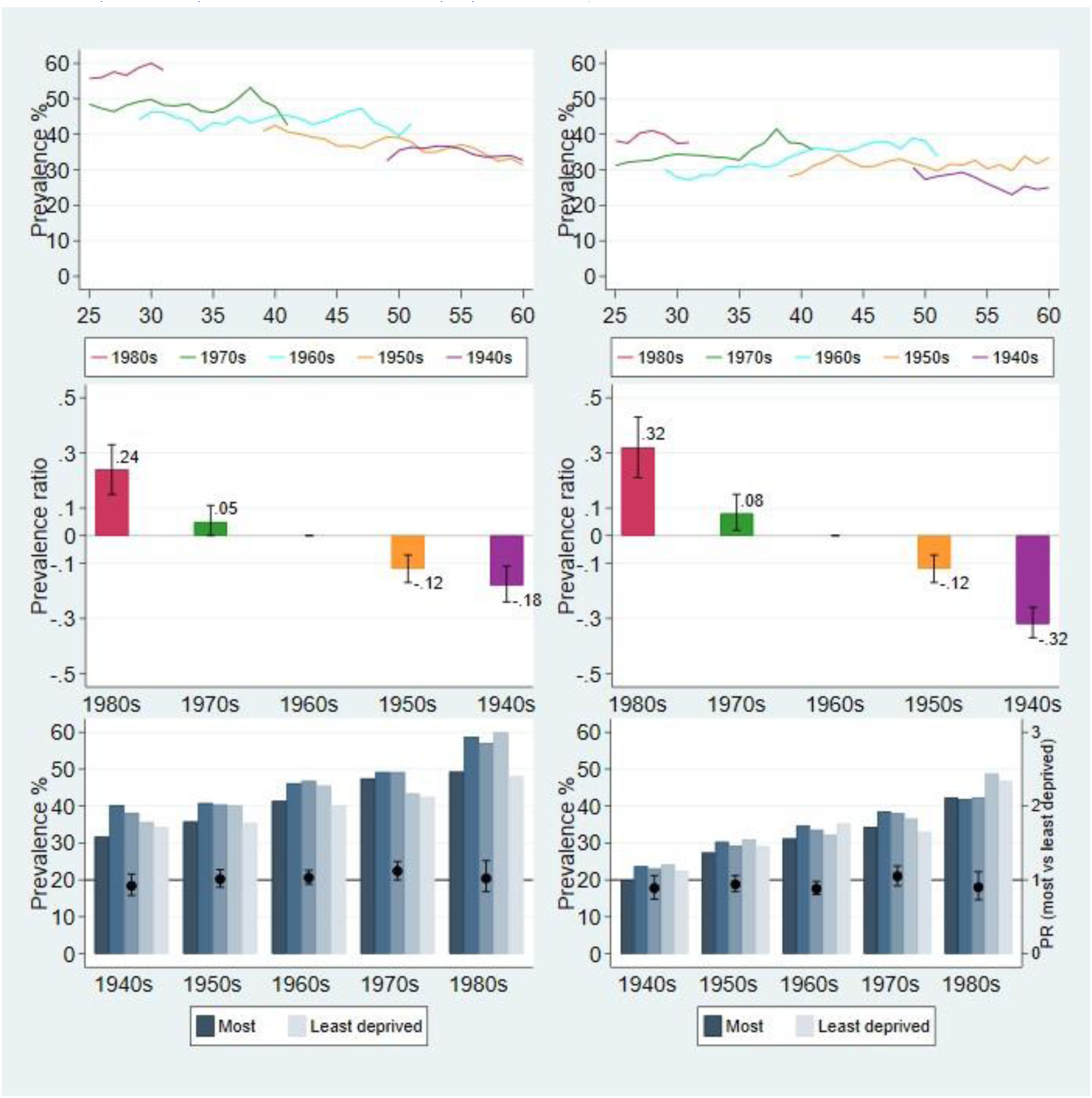
Physically active (spending 30 minutes or more in moderate-or vigorous-intensity activity on at least five days per week) Notes: Males left panel, Females right panel. **Top panel**: 3-year moving averages of observed prevalence (%) by age and cohort. **Middle panel**: Prevalence ratio (value at top of bar), relative to cohort 1960s (rebased to equal 0), with 95% confidence intervals (CIs) **Bottom panel**: Predicted prevalence (%) at age 40, by birth cohort and deprivation quintiles (primary y-axis); prevalence ratio (PR) between most and least deprived quintiles with 95% CIs (secondary y-axis). Unlike top and middle panel, cohorts are plotted from oldest (1940s) to youngest (1980s). PR greater than 1 indicates prevalence is higher in most versus least deprived areas; less than 1 indicates prevalence is lower in most versus least deprived areas. PR is statistically significant at the 95% level when the CIs do not span 1.

After adjustment for age and deprivation, prevalence rates for females were 32% higher for the 1980s cohort and 32% lower for the 1940s cohort (relative to those in the 1960s cohort). For males, the pattern was similar but the gradient between cohorts was less pronounced, with physical activity levels being 24% higher for the 1980s cohort and 18% lower for the 1940s cohort.

#### Inequalities

Physical activity levels increased consistently across all deprivation quintiles, in parallel with cohort progression, with little evidence of inequalities or changes in inequalities (Table 2, S1_Table SD7).

### Fruit and Vegetables consumption

#### Age and cohort differences

Data for fruit and vegetable consumption were collected over a limited number of years in the HSE series (starting from 2001), and the proportion of participants eating five or more portions per day remained relatively constant between the cohorts (Fig 8). There was a consistent age pattern, with individuals aged 25 having slightly lower consumption levels compared to older ages. From approximately age 50 onwards, a greater proportion of females than males reported consuming five or more portions of fruits and vegetables daily.

**Fig 8.**
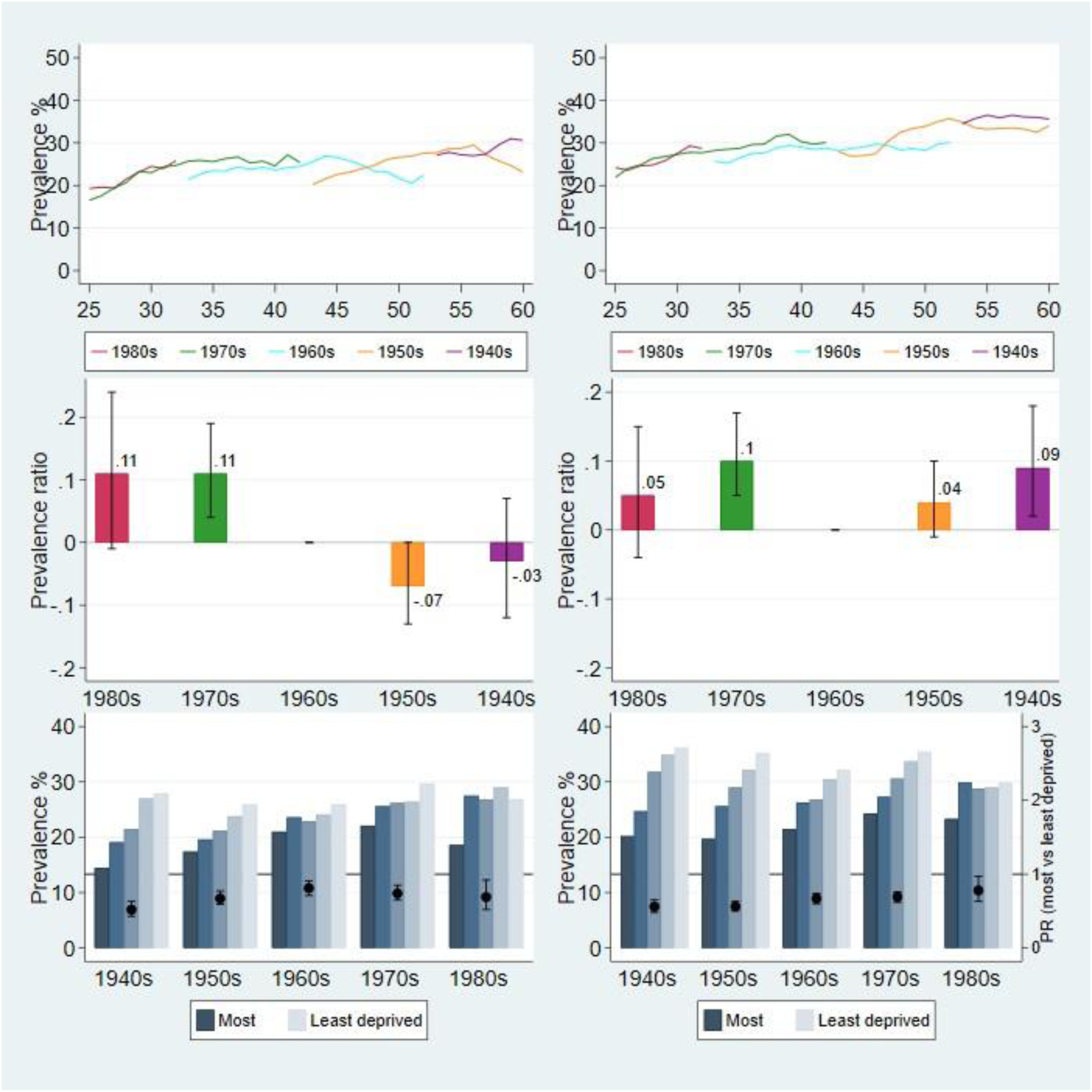
Consumed five or more portions of fruit and vegetables per day. Notes: Males on left, Females on right. **Top panel**: 3-year moving averages of prevalence (%) by age and cohort. **Middle panel**: Prevalence ratio (value at top of bar), relative to cohort 1960s (rebased to equal 0), with 95% confidence intervals (CIs) **Bottom panel**: Predicted prevalence (%) at age 40, by birth cohort and deprivation quintiles (primary y-axis); prevalence ratio (PR) between most and least deprived quintiles with 95% CIs (secondary y-axis). Unlike top and middle panel, cohorts plotted from oldest (1940s) to youngest (1980s). PR greater than 1 indicates prevalence is higher in most versus least deprived areas; less than 1 indicates prevalence is lower in most versus least deprived areas. PR is statistically significant at the 95% level when the CIs do not span 1.

#### Inequalities

Levels of fruit and vegetable consumption at age 40 showed a consistent inequality gradient, with those in the least deprived quintile being more likely to meet the recommended number of portions, compared to their counterparts in the most deprived quintile. However, among both sexes, the absolute and relative gaps in prevalence were lower in the younger versus older cohorts. In males, for example, the PR significantly decreased from 0.52 (95% CI: 0.43-0.63) in the 1940s cohort to 0.74 (95% CI: 0.65-0.85) in the 1970s cohort (Table 2; S1_Table SD8). Likewise, among females, the PR significantly decreased from 0.56 (95% CI: 0.48-0.65) in the 1940s cohort to 0.78 (95% CI: 0.63-0.97) in the 1980s cohort. In simpler terms, the proportion of participants meeting the 5-a-day recommendations increased at a slightly faster pace in the most deprived quintile between successively younger cohorts, though starting from a lower base.

## Discussion

To our knowledge, this study represents the first analysis of generational birth cohort trends which includes five of the seven major risk factors identified by the World Health Organisation to reduce premature mortality from non-communicable diseases by 25% by 2025.^14^ These five risk factors are causally linked to the future burden of morbidity, quality of life and leading causes of premature mortality. Our study maximises the utility of a long running high-quality cross-sectional and nationally-representative health data series by presenting a generational post-war cohort analysis for the population of England at working ages before survival bias substantially distorts patterns observed, particularly for health related analyses. This approach offers an alternative perspective on temporal trends in behavioural risk factor prevalence between cohorts, as well as changes in inequalities within each. Below we discuss our findings for each risk factor in turn.

### Current and heavy smoking

Of the five risk factors studied, our results for current smoking were the most positive, suggesting that age- and deprivation-adjusted prevalence levels fell between the oldest and youngest cohorts, for both sexes. The absolute gap in current smoking prevalence between the most- and least-deprived quintiles narrowed, and relative inequalities fell significantly, for the 1960s cohort compared to the 1940 and 1950 cohorts, and remained at this lower level for subsequent younger cohorts.

The continuous decline in smoking prevalence at working ages in the more recent born cohorts has been attributed mainly to a decline in smoking initiation before the age of 25, and to a lesser extent to smoking cessation thereafter.^15^

These findings align partly with a pair of previous generational studies, despite differences in survey data used and in timeframes.^16, 17^ Basing interpretation on line graphs of observed prevalence, these studies also showed that age-for-age smoking prevalence fell for males and females in younger cohorts, but that the absolute gap between manual and non-manual groups appeared to peak for the 1950s cohort and thereafter stabilised at this higher rate, thereby implying that smoking inequalities might not diminish in more recently born cohorts. In contrast, because we have incorporated data from later rounds of surveys, and used a modelling approach which adjusted for age effects and used a finer gradation of socioeconomic circumstances (by neighbourhood deprivation quintiles), our results showed that both absolute and relative inequalities in smoking prevalence between successive cohorts have fallen markedly.

Our finding of relative inequality reduction from a cohort perspective also appears to be at odds with a recent analysis of temporal trends in socioeconomic inequalities in smoking prevalence between 2003 and 2019 for all adults aged 16 and over.^18^ Using a summary measure of age-standardised smoking prevalence, this study reported an increase in relative inequalities between the most- and least-advantaged groups based on measures of individual socio-economic position. However, similar to our findings, they did not observe a significant increase in either absolute- or relative-inequalities in smoking prevalence when examined by IMD quintiles. It is important to reiterate that annual age-standardised prevalence estimates summarise the average experiences of different combinations of cohorts. This highlights the importance of using different approaches to disentangle any cohort effects on temporal trends, especially when considering what future smoking trends might look like once older cohorts are replaced. In the present study, we specifically examined generational cohort effects to provide an alternative insight into how inequalities in health behaviours have evolved over the last two decades. Besides differences arising due to study design and coverage, messages from different studies can be valid and serve to provide complementary results albeit from a different perspective.

Among current smokers, the prevalence of heavy smoking (20 or more cigarettes a day) markedly decreased across all ages for younger cohorts, and at a similar pace across deprivation quintiles, especially in males. As a result, for males, absolute inequalities at age 40 reduced by about two-thirds from the level observed in the 1940s cohort compared to the 1980s cohort, but with no discernible difference in relative inequalities across cohorts. Females also experienced a sharp decline in absolute inequalities; however, relative inequalities increased over successively younger cohorts, suggesting that females living in the least deprived areas were quicker to reduce consumption compared to their counterparts living in the most deprived areas.

### Alcohol consumption: frequency and amount

With respect to alcohol consumption, longitudinal cohort studies have shown that the pattern of alcohol consumption fluctuates over the life course, with frequency of drinking occasions gradually becoming most prevalent during middle-age, stabilising, and then declining from about age 70.^19^ In the present study, we observed this age-pattern in the cohorts born in the 1960s or earlier. However, in the younger cohorts, this age pattern was absent, and the levels of frequent drinking remained consistently low from age 25 onwards.

The observed between-cohort differences in the frequency of alcohol consumption mirrored that of cigarette smoking, with prevalence falling dramatically for successively younger cohorts, resulting in the narrowing in absolute inequalities. However, unlike smoking, we observed that the social gradient flipped, with participants in both sexes in the least deprived areas showing higher levels of frequent drinking than their counterparts in the most deprived areas across all cohorts. The reversal in drinking habits between more and less advantaged groups connects to the widely reported ‘alcohol harm paradox’, with levels of alcohol related harm and mortality being markedly higher in disadvantaged groups despite lower levels of alcohol consumption.^20^

This paradox may, in part, be explained by the patterns in the prevalence of harmful drinking, defined in the present study as drinking more than twice the daily recommended units on the heaviest drinking day in the past week. Females living in the most deprived quintiles, in particular, consistently had higher rates of harmful drinking compared to all other quintiles. For males, the heavy drinking gradient with deprivation was found to be less clear-cut. Unfortunately, changes in the definition of a standard unit of alcohol in the HSE over the study period limited deeper analysis of differences in daily consumption patterns between birth cohorts and across deprivation quintiles.

Harmful drinking prevalence levels were significantly lower in the 1970s and 1980s cohorts compared to cohorts born in 1960s or older, for both sexes. However, the signal for falls in both absolute and relative inequalities between cohorts was strong from the 1940s until the 1960s cohort, but remained static thereafter.

### Obesity

Temporal trends in obesity among adults have shown a steady increase, with prevalence levels in England approximately doubling between 1993 and 2019 in both sexes.^21^ Obesity levels estimated from HSE data have been shown to be consistently higher in women than in men across all age groups, and higher for the most disadvantaged groups, as indicated by both individual (e.g. equivalised household income quintiles) and area-based measures of socio-economic position. In the present study, as expected, prevalence levels increased monotonically until the age of 60 across all cohorts. Relative to the levels in the 1960s cohort, obesity rates were significantly higher in the 1970s and 1980s cohorts, particularly for females, after adjusting for age and deprivation.

Analyses of long-run trends in inequalities in obesity prevalence are limited. In our study, against a backdrop of rising obesity prevalence, we observed modest inequalities in males in all cohorts. In contrast, the magnitude of absolute inequalities tripled between the older and younger cohorts in females from the 1940s to 1960s cohort, but then stabilised thereafter.

However, the key question underlying the surge in obesity levels across developed countries revolves around the extent to which it can be attributed to period effects (such as increased access in more recent years to inexpensive fast food) and how much to cohort effects (through factors such as sedentary behaviour, and the obesogenic environment). Analyses of obesity patterns conducted using age-period-cohort models in both the United States (using a longitudinal cohort)^22^ and in England (examining generational cohort patterns)^23^ have concluded that cohort effects are just as significant as period effects, especially in more recent cohorts. While the estimates of age-cohort effects presented in our descriptive study incorporate the contribution of period effects, we have not disentangled the independent contributions of each. Reither et al. suggest that birth cohort membership adds an additional risk factor, distinct from period effects that impact all sections of society uniformly.^18^

### Physical activity

The rise in physical activity levels in later-born cohorts observed in our study aligns with secular annual trends showing a year on year increase in physical activity at recommended levels.^24^

Interestingly, across deprivation quintiles, estimated activity levels (presented herein) at age 40 increased at a similar pace across successively younger cohorts, leading to no discernible differences in inequalities. This is in contrast to age-standardised point estimates of differences in physical activity levels for all adults (i.e. aged 16 and over) using HSE 2016 data which showed a graded relationship by IMD quintile, with just over two-thirds of adults living in the least deprived quintile meeting the recommended activity levels as compared to half of those living in the most deprived quintile.^25^ This suggests that age trajectories of activity levels may diverge, following a pattern of decline that varies across different socioeconomic groups as individuals enter older age. Furthermore, differential survival rates between deprivation groups at older age may potentially exacerbate the social gradient observed in prevalence rates estimated among all adults, rather than focusing solely on adults of working age.

### Fruit and vegetables consumption

Statistics using the more accurate food diary collection method in the National Diet and Nutrition Surveys (NDNS) support the findings from the HSE, suggesting minimal change in the proportion of the adult population consuming five or more portions of fruits and vegetables a day from 2001 onwards.^26, 27^ But few studies have analysed point estimates of socioeconomic inequalities in fruit and vegetable consumption, and none have reported on trends in these inequalities over time.

Our analysis by generational cohorts hints at encouraging positive signals in that the absolute and relative gaps in prevalence narrowed in the younger versus older cohorts, reflecting a slightly faster increase in the most deprived areas, albeit from a lower starting base than for those living in the least deprived areas.

Similar to the challenges posed in analysing obesity trends, it is difficult to disentangle the independent contribution of period changes, such as improved availability of fruits and vegetables, from generational cohort differences in consumption influenced by raised awareness of health benefits following the introduction of the 5-a-day programme in the UK in 2003. An analysis of a single British birth cohort (1946 cohort) prospectively showed that the diet of participants became healthier in middle-age. However, as the authors noted, it is impossible to differentiate whether this is an ageing or a period effect.^28^

Our findings suggest that policy actions aimed at promoting awareness of health-enhancing behaviours may have positively influenced the health of successive younger cohorts. Given that the independent relative risks of mortality for cigarette smokers are roughly twice those for obesity, our findings, along with the secular declines in fatal diseases, suggest that cohort life expectancies will likely increase. ^29^ However, this improvement is projected to be offset by higher levels of obesity-related comorbidities and functional disability in later years.^30^

#### Strengths and limitations

To the best of our knowledge this is the first UK generational cohort study to include key modifiable behavioural risk factors causally associated with a range of chronic diseases and all-cause mortality; and to describe both absolute and relative inequalities in these risk factors across post-war generational cohorts of working age. We have quantified the scale of inequality, whether these differences are statistically significant, and estimated the disparities in both absolute and relative dimensions within each cohort.

The source data used in this study are of high quality, with face to face interviews conducted by trained interviewers following standardised protocols and measurement devices. However, other than obesity (BMI calculated from measured height and weight), risk factors were self-reported; thus, measurement errors are inevitable. However, the use of repeated measures of these variables could capture the long-term underlying social change. The continuity of the cross-sectional surveys over 26 years without interruption made it possible to analyse underlying between-cohort change over overlapping ages. Unlike longitudinal studies (of single or mixed-age cohorts), generation-cohort studies are not susceptible to attrition, data collection is less expensive and is more likely to be routinely collected.

A limitation is our focus on cohort change whilst ignoring period effects. The well-known Age-Period-Cohort (APC) identification problem^31^ arising from the perfect correlation between these three dimensions, makes it challenging to separate out the distinct contributions of each to the observed change in behavioural risk factor prevalence without employing more advanced statistical methods. Therefore, it is important to acknowledge that the cohort differences presented herein provide only a *credible* signal of underlying social change which may partly be attributable to period effects. For instance, changes in cigarette smoking within the population could be due, in part, to period measures such as indoor smoking bans (reducing smoking initiation in younger cohorts) or the increased availability of smoking cessation services (leading to a steeper decline with advancing age in successively younger cohorts).

Secondly, despite the large size of the pooled dataset, the scope of our analysis for some risk factors was sometimes restricted by breaks in data collection or changes in definition, adding to the small data problem once the analytic sample was stratified by age, sex, area deprivation and cohort. In addition, the constructed cohorts did not cover the same age ranges. We tried two methods to overcome this: age standardisation and a model-based approach. Using the 1960s cohort as reference (since it overlapped the majority of the entire study age range), we compared the directly age-standardised prevalence rates of the age-truncated cohorts against the corresponding rates for the 1960s cohort (results available on request). The age-adjusted prevalence ratios we obtained from the two methods were found to be very similar, providing further confidence in our opting to report results based on the model-based approach.

In our analysis of inequalities within- and between-cohorts, we only included the main effect of age, and we focused our presentation on the estimates of inequalities at age 40, the midpoint of our pooled dataset. However, it is worth noting that socioeconomic inequalities at other ages may exhibit different patterns. Inequalities could be more pronounced at younger ages due to factors such as the most advantaged groups having lower rates of smoking initiation or a more rapid pace of smoking cessation with age. The choice to present inequalities at age 40 was a practical decision for our analysis, but we acknowledge that exploring variations at different ages could provide additional insights into the dynamics of these inequalities in the working age population.

## Conclusions

Our analysis of generational change reveals credible signals of behavioural risk factor changes in levels and in inequalities over successive birth cohorts of working age. In terms of absolute inequality, there is a positive health signal as smoking prevalence and alcohol consumption have reduced, while physical activity has increased across all deprivation groups. However, we observe a concerning negative health signal regarding obesity prevalence, particularly in females, which has increased for all socioeconomic groups across successively younger cohorts. These findings underscore the complex interplay of factors influencing health disparities and behaviours among different socioeconomic groups over time.

## Acknowledgments

The authors thank the interviewers and nurses, and participants of the Health Surveys for England 1994-2019. NatCen Social Research, and NHS England are responsible for designing, collecting and administering the survey data and making the data available for public use via the UK Data Service.

We thank Dr Zander Crook (left UCL in 2022) for compiling the pooled survey dataset, 1994-2019.

## Contributions

MB conceptualised, designed and supervised the study. SS led on the statistical modelling and advised on behavioural risk factor definitions. MB and SS were jointly responsible for conducting the analyses (including graphical presentation) and interpreting the results. MB drafted the manuscript, with input from SS on statistical methods and risk factor definitions. Both authors have read and approved the manuscript. MB is responsible for the overall content as guarantor.

## Funding

MB is honorary research staff at UCL, funded by Legal & General Assurance Society Limited (L&G) as part of its wider research collaboration with UCL on longevity research. SS received no specific funding for this work. The funders had no role in study design, data collection and analysis, decision to publish, or preparation of the manuscript.

## Competing Interests

MB has support from Legal and General Assurance Society Limited for the submitted work; SS has no relationship with Legal and General Assurance Society Limited in the submitted work or in the previous 3 years. The authors have no other competing interests.

## Data availability

Data are available in a public, open access repository. The HSE datasets generated and analysed during the current study (age banding for participants) are available via the UK Data Service (UKDS: https://ukdataservice.ac.uk/), subject to their end user license agreement.

## Ethical approval and participant consent

In the HSE, all adults in selected households are eligible for interview. Relevant committees granted research ethics approval for the survey. Participants gave verbal consent for interview.

## References

1 Bajekal M, Scholes S, Love H, et al. Analysing recent socioeconomic trends in coronary heart disease mortality in England, 2000-2007: a population modelling study. PLoS Med 2012;9:e1001237. doi: 10.1371/journal.pmed.1001237.

2 Scholes S, Bajekal M, Love H, et al. Persistent socioeconomic inequalities in cardiovascular risk factors in England over 1994-2008: a time-trend analysis of repeated cross-sectional data. BMC Public Health 2012;12:129. doi: 10.1186/1471-2458-12-129.

3 Deaton, Angus. The analysis of household surveys: a microeconometric approach to development policy. 1997 © Washington, DC: World Bank. http://hdl.handle.net/10986/30394 License: CC BY 3.0 IGO.

4 Li Y, Pan A, Wang DD et al. Impact of Healthy Lifestyle Factors on Life Expectancies in the US Population. Circulation 2018;138:345–355. doi: 10.1161/CIRCULATIONAHA.117.032047.

5 Stringhini S, Dugravot A, Shipley M, et al. Health Behaviours, Socioeconomic Status, and Mortality: Further Analyses of the British Whitehall II and the French GAZEL Prospective Cohorts. PLoS Med 2011;8:e1000419. doi:10.1371/journal.pmed.1000419

6 Marteau T, Rutter H, Marmot M. Changing behaviour: an essential component of tackling health inequalities. BMJ 2021;372:n332.

7 Mindell J, Biddulph JP, Hirani V et al. Cohort profile: the Health Survey for England. International Journal of Epidemiology 2012;41:1585–1593. doi:10.1093/ije/dyr199.

8 Noble M, Mclennan D, Wilkinson A, Whitworth S, Exley H. Barnes, and Dibben C. (2007). The English indices of deprivation 2007. Department of Communities and Local Government.

9 Lloyd, CD, Norman, PD & McLennan, D. Deprivation in England, 1971–2020. Appl. Spatial Analysis 2023;16:461–484. 10.1007/s12061-022-09486-8

10. Macintyre S, Ellaway A, Cummins S. Place effects on health. How do we conceptualise, operationalise and measure them? Soc Sci Med 2002;55:125–39. doi: 10.1016/s0277-9536(01)00214-3.PMID: 12137182

11. Barros AJ, Hirakata VN. Alternatives for logistic regression in cross-sectional studies: an empirical comparison of models that directly estimate the prevalence ratio. BMC Med Res Methodol 2003;3:21. doi: 10.1186/1471-2288-3-21.

12 Jivraj, S. Are self-reported health inequalities widening by income? An analysis of British pseudo birth cohorts born, 1920-1970. J Epidemiol Community Health 2020;74:255–259. doi: 10.1136/jech-2019-213186.

13 Jivraj S, Goodman A, Pongiglione B, Ploubidis GB. Living longer but not necessarily healthier: The joint progress of health and mortality in the working-age population of England. Popul Stud (Camb) 2020;74:399–414. doi:10.1080/00324728.2020.1767297.

14 Stringhini S, Carmeli C, Jokela M, et al. Socioeconomic status and the 25 × 25 risk factors as determinants of premature mortality: a multicohort study and meta-analysis of 1·7 million men and women. Lancet 2017;389:1229–1237. doi: 10.1016/S0140-6736(16)32380-7.

15 Opazo Breton M, Gillespie D, Pryce R, et al. Understanding long-term trends in smoking in England, 1972-2019: an age-period-cohort approach. Addiction 2022;117:1392–1403. doi: 10.1111/add.15696. Epub 2021 Oct 21.

16. Davy M. Time and generational trends in smoking among men and women in Great Britain, 1972-2004/05. Health Stat Q 2006;32:35–43.

17 Davy M. Socio-economic inequalities in smoking: an examination of generational trends in Great Britain. Health Stat Q. 2007;34:26–34.

18 Ogunlayi F, Coleman PC, Fat LN, et al. Trends in socioeconomic inequalities in behavioural non-communicable disease risk factors: analysis of repeated cross-sectional health surveys in England between 2003 and 2019. BMC Public Health 2023;23:1442. doi: 10.1186/s12889-023-16275-6.

19 Britton A, Ben-Shlomo Y, Benzeval M, et al. Life course trajectories of alcohol consumption in the United Kingdom using longitudinal data from nine cohort studies. BMC Med 2015;13:47. doi: 10.1186/s12916-015-0273-z.

20 Bellis MA, Hughes K, Nicholls J, Sheron N, Gilmore I, Jones L. The alcohol harm paradox: using a national survey to explore how alcohol may disproportionately impact health in deprived individuals. BMC Public Health. 2016;16:111. doi: 10.1186/s12889-016-2766-x.

21 Patterns and trends in obesity, Public Health Analysis Directorate, Office for Health Improvement and Disparities. 26.02.2020.

22 Reither EN, Hauser RM, Yang Y. Do birth cohorts matter? Age-period-cohort analyses of the obesity epidemic in the United States. Soc Sci Med 69;10:1439–48. doi: 10.1016/j.socscimed.2009.08.040.

23 Opazo Breton M, Gray LA. An age-period-cohort approach to studying long-term trends in obesity and overweight in England (1992-2019). Obesity (Silver Spring). 2023;31:823–831. doi: 10.1002/oby.23657.

24 Natcen, UCL. Adult health trends. Health Survey for England 2016. Available at: HSE 2016 Adult report (digital.nhs.uk).

25 Scholes, S. Physical activity in adults. Health Survey for England 2016. NHS Digital. Available at: hse16-adult-phy-act.pdf (digital.nhs.uk).

26 Whitton C, Nicholson SK, Roberts C, et al. National Diet and Nutrition Survey: UK food consumption and nutrient intakes from the first year of the rolling programme and comparisons with previous surveys. Br J Nutr 2011;106:1899–914. doi: 10.1017/S0007114511002340.

27. Public Health England. NDNS: results from years 9 to 11 (combined) – statistical summary - GOV.UK (www.gov.uk). 2020. Available at: https://www.gov.uk/government/statistics/ndns-results-from-years-9-to-11-2016-to-2017-and-2018-to-2019/ndns-results-from-years-9-to-11-combined-statistical-summary.

28 Pot GK, Prynne CJ, Almoosawi S, NSHD scientific and data collection teams. Trends in food consumption over 30 years: evidence from a British birth cohort. Eur J Clin Nutr 2015;69:817–23. doi: 10.1038/ejcn.2014.223.

29 Stringhini S, Carmeli C, Jokela M, et al. Socioeconomic status and the 25 × 25 risk factors as determinants of premature mortality: a multicohort study and meta-analysis of 1·7 million men and women. Lancet 2017;389:1229–1237. doi: 10.1016/S0140-6736(16)32380-7.

30. The Health Foundation. Health in 2040: projected pattern of illness in England. Insight report, July 2023. https://www.health.org.uk/publications/health-in-2040.

31 Bell A. (2020). Age period cohort analysis: a review of what we should and shouldn’t do. Ann Hum Biol. 2020;47:208–217. doi: 10.1080/03014460.2019.1707872.

