## Supplementary material for "Inequalities in behavioural risk factor prevalence between five post-war generational cohorts of working age, England": S1_Supplementary Appendix: Bajekal Scholes_Generational change _S1 Supplementary.pdf

### S1. Supplementary Appendix for 'Differentials in behavioural risk factors between five post-war generational cohorts of working age, England'

- Section A. Risk factor variables definitions
- Section B. Sample characteristics and distribution
- Section C. Tables underlying observed cohort prevalence rates for risk factors
- Section D. Tables of predicted prevalence and prevalence ratios by Index of Multiple Deprivation (IMD) quintiles at age 40 (absolute and relative inequalities)

#### A: Risk factor variables definitions.

**Self-reported current smoking status**

Two binary variables were derived: percent of those who were current cigarette smokers and, of smokers, those who were classified as heavy smokers (smoking 20 or more cigarettes per day).

Questions on smoking were asked each year. Participants were asked whether they had ever smoked a cigarette, and those who reported having ever smoked were asked whether they smoked cigarettes at all nowadays. In this study participants were classified either as current smokers or not current smokers (i.e. ex-/never-smokers).

Participants who reported smoking nowadays were also asked about the number of cigarettes smoked daily (on weekdays and at weekends). Responses were grouped into 1 = Light smokers, under 10 a day; 2 = Moderate smokers, 10 to under 20 a day; 3 = Heavy smokers, 20 or more a day; 4 = Don't know number smoked a day; 5 = Non-smoker. Data on smoking intensity were not available by single year of age in the End User Licence (EUL) datasets available at the UK Data Service for years between 2014-2019, and so these years have not been included in our analyses.

**Alcohol**

Two binary variable were derived: alcohol consumption frequency (to assess usual pattern of drinking in preceding last 12 months); and, of those reported having drunk alcohol last week, the percentage defined as 'heavy drinkers'.

Questions on alcohol were asked each year, but with differences in the questions asked which affected the number of years of data we could use to define a consistent series. Questions cover how often participants had an alcoholic drink of any kind during the last 12 months (ranging from almost every day to not at all). A high usual pattern of alcohol consumption was defined in this study as drinking alcohol on five or more days per week.

Alcohol consumption (type and quantity) on the heaviest drinking day in the last week was asked in all years. However, the measure of units of alcohol was changed in 2007 and hence for consistency over time, we have derived estimates of heavy drinking from 2007 onwards only in this analysis

(Fuller 2007). Of those who reported having drunk alcohol at all last week, we defined 'heavy drinkers' as, of those who had drunk last week, the proportion who consumed 8 or more units for men and 6 or more units for women on the heaviest drinking day last week (ONS 2002).

**Obesity**

BMI was calculated as weight in kilograms (kg) divided by height in metres squared. Obesity was defined as BMI 30 kg/m$^2$ or more.

Height and weight were measured by trained interviewers each year. Height was measured using a portable stadiometer with a sliding head plate, a base plate and connecting rods marked with a measuring scale. Participants were asked to remove their shoes. One measurement (to the nearest even millimetre) was taken, with participants' stretching to the maximum height and the head positioned in the Frankfort plane. For those not pregnant, a single weight measurement (to the nearest 100 g; maximum 130kg [1994-2011] and maximum 200kg [2012 onwards]) was recorded using Class III Seca scales; participants were asked to remove their shoes and any bulky clothing or heavy items in pockets, etc. No adjustment was made for the weight of clothing. Participants unable to stand or unsteady on their feet were not measured. Participants were assigned missing values if they were considered by the interviewer to have unreliable measurements, for example, those who were too stooped or wore excessive clothing.

**Physical activity**

A high level of physical activity was defined in this study as spending 30 minutes or more in moderate- or vigorous-intensity activity on at least five days per week.

Questions on physical activity assess frequency (number of days spent doing a specified activity in the last four weeks) and duration (of an average episode/bout lasting above a specified duration limit) in four leisure-time domains: domestic activity, DIY/manual work, walking, and sports/exercise. Physical activity undertaken whilst at work was also taken into account in the estimation of summary activity levels. Physical activities have been classified into intensity levels (light, moderate, vigorous) based on an estimate of the energy cost of the activities.

Changes in the physical activity questions have restricted the meaningfulness of comparisons over time to some extent. Full details are provided elsewhere (Scholes and Mindell 2013). For example, the lower duration limit for an activity to be included was 15 minutes in 1998 and 2006, 30 minutes in 2003 (15 minutes for sports and exercise), and 10 minutes in 2008, 2012, and 2016. A single question on occupational physical activity ('Thinking about your job, in general would you say that you are very physically active, fairly physically active, not very physically active, or not at all physically active?') was asked in 2003 and 2006; more detailed questions introduced in 2008 focus on what people actually do at work, and how many hours they typically work.

To maximise the number of data points, we derived a variable summarising the number of days per week that participants undertook physical activity of at least moderate intensity for a minimum duration of 30 minutes. Based on the single question relating to occupational physical activity, for those participants who reported that they were very or fairly active in their job, arbitrary estimates of 12 or 20 working days (3 or 5 days/week respectively) were used, depending on whether the participant worked part- or full-time, to assess levels of physical activity whilst at work.

Data on adult physical activity were not available by single year of age in the dataset available via EUL from the UK Data Service for 2016 when this module was included in the HSE, and so 2016 data has not been included in our analyses.

**Fruit and vegetable consumption**

Healthy eating was defined as consuming five or more portions of fruits or vegetables per day.

Since 2001 (but not in 2012 and 2014), participants have been asked about fruit and vegetable consumption on the previous day (the 24 hours from midnight to midnight). Participants are asked about all vegetables and fresh, canned and frozen fruit, as well as salad, pulses, dried fruit and fruit juice/smoothies they have consumed, including dishes made mainly from fruit or vegetables ('composites'). Participants' responses are coded into portion sizes. A maximum of one portion of pulses, one of fruit juice or a smoothie, and one of dried fruit can contribute to the total daily portions of fruit and vegetables.

Data for 2014-2019 were not available by single year of age in the EUL datasets available via the UK Data Service and so have not been included in the analyses.

References

Fuller, E. Alcohol consumption in Craig R and Mindell J (eds), 2007. Health Survey for England 2006 Cardiovascular disease and risk factors in adults: vol 1, NHS Information Centre 2007.

https://files.digital.nhs.uk/publicationimport/pub01xxx/pub01213/heal-surv-cvd-risk-obes-ad-ch-eng-2006-rep-v1.pdf

Office for National Statistics (ONS), 2002. Living in Britain: Results from the 2001 General Household Survey. The Stationery Office, London.

Scholes, S and Mindell J. Physical activity in adults in Craig R and Mindell J (eds). Health Survey for England 2012 Health, social care and lifestyles: vol 1, The Health and Social Care Information Centre 2013. https://files.digital.nhs.uk/publicationimport/pub13xxx/pub13218/hse2012-ch2-phys-act-adults.pdf

#### B. Sample characteristics and distribution

SB.1: Pooled sample size, by birth cohort and age (persons)

| AGE | 1940S | 1950S | 1960S | 1970S | 1980S | OVERALL |
|---|---|---|---|---|---|---|
| **60** | 1617 | 1284 | | | | |
| **59** | 1669 | 1190 | 113 | | | |
| **58** | 1635 | 1337 | 279 | | | |
| **57** | 1695 | 1350 | 464 | | | |
| **56** | 1683 | 1335 | 578 | | | |
| **55** | 1835 | 1479 | 667 | | | |
| **54** | 1969 | 1395 | 895 | | | |
| **53** | 1757 | 1473 | 990 | | | |
| **52** | 1697 | 1544 | 1202 | | | |
| **51** | 1535 | 1615 | 1265 | | | |
| **50** | 1448 | 1689 | 1398 | | | |
| **49** | 1298 | 1706 | 1351 | 122 | | |
| **48** | 1028 | 1728 | 1513 | 276 | | |
| **47** | 914 | 1765 | 1500 | 412 | | |
| **46** | 538 | 1732 | 1660 | 552 | | |
| **45** | 290 | 2009 | 1616 | 720 | | |
| **44** | | 2184 | 1692 | 834 | | |
| **43** | | 1911 | 1714 | 987 | | |
| **42** | | 1758 | 1837 | 1108 | | |
| **41** | | 1588 | 2008 | 1189 | | |
| **40** | | 1413 | 1977 | 1375 | | |
| **39** | | 1338 | 2042 | 1310 | 143 | |
| **38** | | 1020 | 2136 | 1469 | 279 | |
| **37** | | 942 | 2105 | 1379 | 417 | |
| **36** | | 617 | 2127 | 1484 | 527 | |
| **35** | | 345 | 2334 | 1468 | 677 | |
| **34** | | | 2554 | 1501 | 797 | |
| **33** | | | 2224 | 1574 | 943 | |
| **32** | | | 2091 | 1487 | 1026 | |
| **31** | | | 1781 | 1545 | 1130 | |
| **30** | | | 1639 | 1563 | 1194 | |
| **29** | | | 1446 | 1603 | 1114 | |
| **28** | | | 1109 | 1598 | 1193 | |
| **27** | | | 883 | 1543 | 1147 | |
| **26** | | | 567 | 1551 | 1137 | |
| **25** | | | 281 | 1666 | 1139 | |
| ***TOTAL N*** | **22,608** | **37,747** | **50,038** | **30,316** | **12,863** | **153,572** |
| **MEAN** | 1413 | 1452 | 1430 | 1213 | 858 | 1313 |
| **MEDIAN** | 1626 | 1476 | 1513 | 1468 | 1026 | 1413 |

SB.2 Pooled sample distribution by birth cohort and IMD deprivation quintiles (persons)

| | LEAST DEPRIVED | 2.00 | 3.00 | 4.00 | MOST DEPRIVED | MISSING | TOTAL BY COHORT |
|---|---|---|---|---|---|---|---|
| **1940S (N)** | 5065 | 4762 | 4305 | 3542 | 3069 | 1865 | 22608 |
| **ROW %** | *22.4* | *21.1* | *19.0* | *15.7* | *13.6* | *8.2* | *14.7* |
| **1950S (N)** | 8316 | 7501 | 7149 | 6369 | 5587 | 2825 | 37747 |
| **ROW %** | *22.0* | *19.9* | *18.9* | *16.9* | *14.8* | *7.5* | *24.6* |
| **1960S (N)** | 9921 | 9518 | 9396 | 9107 | 8591 | 3505 | 50038 |
| **ROW %** | *19.8* | *19.0* | *18.8* | *18.2* | *17.2* | *7.0* | *32.6* |
| **1970S (N)** | 5413 | 5230 | 5613 | 5973 | 5826 | 2261 | 30316 |
| **ROW %** | *17.9* | *17.3* | *18.5* | *19.7* | *19.2* | *7.5* | *19.7* |
| **1980S (N)** | 1828 | 2098 | 2530 | 2846 | 3240 | 321 | 12863 |
| **ROW %** | *14.2* | *16.3* | *19.7* | *22.1* | *25.2* | *2.5* | *8.4* |
| **TOTAL BY IMD** | 30543 | 29109 | 28993 | 27837 | 26313 | 10777 | 153572 |
| | *19.9* | *19.0* | *18.9* | *18.1* | *17.1* | *7.0* | *100.0* |
| **% EXCLUDING MISSING** | ***20.6*** | ***19.7*** | ***19.6*** | ***18.8*** | ***17.8*** | | |

#### C: Tables underlying observed cohort prevalence rates for risk factors

Table SC.1. Proportion **current smokers**, by sex, birth cohort and age (3-year moving averages).

| | Males | | | | | Females | | | | |
|---|---|---|---|---|---|---|---|---|---|---|
| **Age** | **1940s** | **1950s** | **1960s** | **1970s** | **1980s** | **1940s** | **1950s** | **1960s** | **1970s** | **1980s** |
| 25 | | | | 0.42 | 0.37 | | | | 0.37 | 0.30 |
| 26 | | | 0.39 | 0.40 | 0.35 | | | 0.38 | 0.34 | 0.30 |
| 27 | | | 0.38 | 0.39 | 0.34 | | | 0.37 | 0.32 | 0.28 |
| 28 | | | 0.38 | 0.39 | 0.33 | | | 0.35 | 0.31 | 0.26 |
| 29 | | | 0.38 | 0.37 | 0.32 | | | 0.34 | 0.29 | 0.24 |
| 30 | | | 0.37 | 0.36 | 0.29 | | | 0.32 | 0.28 | 0.22 |
| 31 | | | 0.38 | 0.35 | 0.28 | | | 0.31 | 0.26 | 0.22 |
| 32 | | | 0.36 | 0.34 | 0.28 | | | 0.30 | 0.26 | 0.20 |
| 33 | | | 0.35 | 0.32 | 0.27 | | | 0.30 | 0.25 | 0.18 |
| 34 | | | 0.33 | 0.32 | 0.25 | | | 0.30 | 0.24 | 0.18 |
| 35 | | | 0.33 | 0.30 | 0.22 | | | 0.30 | 0.23 | 0.17 |
| 36 | | 0.32 | 0.32 | 0.29 | 0.20 | | 0.30 | 0.30 | 0.22 | 0.17 |
| 37 | | 0.32 | 0.31 | 0.26 | 0.20 | | 0.29 | 0.29 | 0.21 | 0.15 |
| 38 | | 0.32 | 0.30 | 0.26 | 0.19 | | 0.28 | 0.28 | 0.20 | 0.16 |
| 39 | | 0.32 | 0.30 | 0.23 | | | 0.28 | 0.27 | 0.19 | |
| 40 | | 0.32 | 0.31 | 0.23 | | | 0.29 | 0.26 | 0.19 | |
| 41 | | 0.31 | 0.29 | 0.22 | | | 0.29 | 0.26 | 0.19 | |
| 42 | | 0.31 | 0.27 | 0.22 | | | 0.29 | 0.25 | 0.19 | |
| 43 | | 0.30 | 0.26 | 0.21 | | | 0.29 | 0.24 | 0.17 | |
| 44 | | 0.30 | 0.24 | 0.23 | | | 0.29 | 0.23 | 0.18 | |
| 45 | | 0.29 | 0.24 | 0.22 | | | 0.29 | 0.22 | 0.17 | |
| 46 | 0.31 | 0.29 | 0.23 | 0.22 | | 0.26 | 0.28 | 0.21 | 0.17 | |
| 47 | 0.30 | 0.29 | 0.23 | 0.21 | | 0.27 | 0.28 | 0.20 | 0.15 | |
| 48 | 0.30 | 0.28 | 0.23 | 0.20 | | 0.29 | 0.27 | 0.20 | 0.15 | |
| 49 | 0.28 | 0.26 | 0.23 | | | 0.29 | 0.25 | 0.20 | | |
| 50 | 0.29 | 0.25 | 0.23 | | | 0.28 | 0.24 | 0.20 | | |
| 51 | 0.28 | 0.25 | 0.22 | | | 0.28 | 0.22 | 0.19 | | |
| 52 | 0.27 | 0.25 | 0.22 | | | 0.28 | 0.22 | 0.18 | | |
| 53 | 0.26 | 0.24 | 0.21 | | | 0.28 | 0.23 | 0.18 | | |
| 54 | 0.25 | 0.24 | 0.22 | | | 0.27 | 0.22 | 0.18 | | |
| 55 | 0.24 | 0.23 | 0.21 | | | 0.25 | 0.21 | 0.16 | | |
| 56 | 0.24 | 0.20 | 0.18 | | | 0.23 | 0.20 | 0.16 | | |
| 57 | 0.24 | 0.18 | 0.18 | | | 0.23 | 0.19 | 0.15 | | |
| 58 | 0.24 | 0.19 | 0.16 | | | 0.21 | 0.19 | 0.16 | | |
| 59 | 0.22 | 0.18 | | | | 0.20 | 0.16 | | | |
| 60 | 0.21 | 0.17 | | | | 0.19 | 0.15 | | | |

Table SC.2. Of current smokers, **heavy smokers** (smoking 20 or more cigarettes per day), by sex, birth cohort and age (3-year moving averages).

| | Males | | | | | Females | | | | |
|---|---|---|---|---|---|---|---|---|---|---|
| **Age** | **1940s** | **1950s** | **1960s** | **1970s** | **1980s** | **1940s** | **1950s** | **1960s** | **1970s** | **1980s** |
| 25 | | | | 0.24 | 0.18 | | | | 0.19 | 0.10 |
| 26 | | | 0.27 | 0.24 | 0.18 | | | 0.19 | 0.20 | 0.12 |
| 27 | | | 0.28 | 0.25 | 0.15 | | | 0.22 | 0.20 | 0.13 |
| 28 | | | 0.30 | 0.24 | 0.15 | | | 0.25 | 0.20 | 0.14 |
| 29 | | | 0.31 | 0.24 | 0.16 | | | 0.29 | 0.21 | 0.13 |
| 30 | | | 0.34 | 0.22 | 0.16 | | | 0.27 | 0.22 | 0.14 |
| 31 | | | 0.37 | 0.23 | 0.12 | | | 0.28 | 0.22 | 0.11 |
| 32 | | | 0.37 | 0.22 | 0.14 | | | 0.28 | 0.21 | 0.15 |
| 33 | | | 0.37 | 0.23 | | | | 0.30 | 0.18 | |
| 34 | | | 0.36 | 0.24 | | | | 0.29 | 0.19 | |
| 35 | | | 0.37 | 0.26 | | | | 0.28 | 0.19 | |
| 36 | | 0.41 | 0.37 | 0.29 | | | 0.36 | 0.29 | 0.20 | |
| 37 | | 0.41 | 0.37 | 0.28 | | | 0.35 | 0.30 | 0.19 | |
| 38 | | 0.44 | 0.36 | 0.27 | | | 0.36 | 0.32 | 0.18 | |
| 39 | | 0.46 | 0.36 | 0.23 | | | 0.39 | 0.32 | 0.17 | |
| 40 | | 0.48 | 0.35 | 0.21 | | | 0.36 | 0.32 | 0.18 | |
| 41 | | 0.47 | 0.38 | 0.24 | | | 0.39 | 0.29 | 0.21 | |
| 42 | | 0.49 | 0.34 | 0.27 | | | 0.37 | 0.30 | 0.18 | |
| 43 | | 0.48 | 0.33 | | | | 0.36 | 0.27 | | |
| 44 | | 0.51 | 0.31 | | | | 0.37 | 0.26 | | |
| 45 | | 0.50 | 0.34 | | | | 0.38 | 0.23 | | |
| 46 | 0.50 | 0.48 | 0.36 | | | 0.37 | 0.38 | 0.26 | | |
| 47 | 0.50 | 0.45 | 0.36 | | | 0.37 | 0.35 | 0.25 | | |
| 48 | 0.55 | 0.46 | 0.34 | | | 0.34 | 0.34 | 0.26 | | |
| 49 | 0.54 | 0.46 | 0.34 | | | 0.36 | 0.36 | 0.22 | | |
| 50 | 0.54 | 0.45 | 0.35 | | | 0.37 | 0.35 | 0.24 | | |
| 51 | 0.52 | 0.44 | 0.31 | | | 0.41 | 0.36 | 0.23 | | |
| 52 | 0.55 | 0.46 | 0.28 | | | 0.41 | 0.30 | 0.27 | | |
| 53 | 0.55 | 0.46 | | | | 0.39 | 0.28 | | | |
| 54 | 0.54 | 0.44 | | | | 0.38 | 0.26 | | | |
| 55 | 0.51 | 0.42 | | | | 0.38 | 0.27 | | | |
| 56 | 0.51 | 0.42 | | | | 0.38 | 0.27 | | | |
| 57 | 0.48 | 0.42 | | | | 0.36 | 0.27 | | | |
| 58 | 0.45 | 0.45 | | | | 0.35 | 0.27 | | | |
| 59 | 0.42 | 0.39 | | | | 0.33 | 0.30 | | | |
| 60 | 0.43 | 0.41 | | | | 0.33 | 0.30 | | | |

Table SC.3. Proportion drinking alcohol on 5 or more days per week, by sex, birth cohort and age (3-year moving averages).

| | Males | | | | | Females | | | | |
|---|---|---|---|---|---|---|---|---|---|---|
| **Age** | **1940s** | **1950s** | **1960s** | **1970s** | **1980s** | **1940s** | **1950s** | **1960s** | **1970s** | **1980s** |
| 25 | | | | 0.17 | 0.09 | | | | 0.08 | 0.04 |
| 26 | | | 0.19 | 0.17 | 0.09 | | | 0.06 | 0.08 | 0.03 |
| 27 | | | 0.18 | 0.17 | 0.08 | | | 0.07 | 0.08 | 0.03 |
| 28 | | | 0.18 | 0.15 | 0.07 | | | 0.08 | 0.08 | 0.03 |
| 29 | | | 0.18 | 0.15 | 0.07 | | | 0.09 | 0.08 | 0.03 |
| 30 | | | 0.18 | 0.14 | 0.07 | | | 0.10 | 0.08 | 0.03 |
| 31 | | | 0.19 | 0.15 | 0.08 | | | 0.10 | 0.08 | 0.03 |
| 32 | | | 0.19 | 0.15 | 0.08 | | | 0.12 | 0.08 | 0.03 |
| 33 | | | 0.20 | 0.14 | 0.11 | | | 0.11 | 0.07 | 0.04 |
| 34 | | | 0.21 | 0.14 | 0.11 | | | 0.11 | 0.07 | 0.03 |
| 35 | | | 0.21 | 0.13 | 0.10 | | | 0.12 | 0.07 | 0.04 |
| 36 | | 0.19 | 0.22 | 0.13 | 0.09 | | 0.13 | 0.13 | 0.07 | 0.04 |
| 37 | | 0.21 | 0.22 | 0.12 | 0.07 | | 0.13 | 0.14 | 0.07 | 0.04 |
| 38 | | 0.23 | 0.22 | 0.12 | 0.06 | | 0.14 | 0.13 | 0.08 | 0.06 |
| 39 | | 0.25 | 0.20 | 0.12 | | | 0.16 | 0.13 | 0.07 | |
| 40 | | 0.26 | 0.21 | 0.13 | | | 0.15 | 0.14 | 0.07 | |
| 41 | | 0.27 | 0.21 | 0.13 | | | 0.17 | 0.14 | 0.06 | |
| 42 | | 0.28 | 0.21 | 0.12 | | | 0.16 | 0.14 | 0.06 | |
| 43 | | 0.28 | 0.20 | 0.10 | | | 0.17 | 0.13 | 0.06 | |
| 44 | | 0.28 | 0.20 | 0.12 | | | 0.17 | 0.13 | 0.06 | |
| 45 | | 0.28 | 0.20 | 0.12 | | | 0.18 | 0.13 | 0.06 | |
| 46 | 0.27 | 0.28 | 0.20 | 0.12 | | 0.17 | 0.18 | 0.13 | 0.06 | |
| 47 | 0.28 | 0.29 | 0.19 | 0.11 | | 0.18 | 0.17 | 0.12 | 0.06 | |
| 48 | 0.29 | 0.28 | 0.17 | 0.13 | | 0.19 | 0.17 | 0.12 | 0.08 | |
| 49 | 0.30 | 0.28 | 0.17 | | | 0.19 | 0.18 | 0.11 | | |
| 50 | 0.32 | 0.26 | 0.18 | | | 0.20 | 0.18 | 0.12 | | |
| 51 | 0.33 | 0.26 | 0.19 | | | 0.20 | 0.18 | 0.12 | | |
| 52 | 0.34 | 0.25 | 0.18 | | | 0.19 | 0.17 | 0.12 | | |
| 53 | 0.32 | 0.25 | 0.17 | | | 0.18 | 0.18 | 0.11 | | |
| 54 | 0.32 | 0.26 | 0.16 | | | 0.19 | 0.18 | 0.11 | | |
| 55 | 0.32 | 0.26 | 0.19 | | | 0.19 | 0.17 | 0.12 | | |
| 56 | 0.33 | 0.25 | 0.21 | | | 0.20 | 0.16 | 0.11 | | |
| 57 | 0.33 | 0.24 | 0.22 | | | 0.19 | 0.16 | 0.11 | | |
| 58 | 0.32 | 0.24 | 0.19 | | | 0.19 | 0.16 | 0.12 | | |
| 59 | 0.33 | 0.24 | | | | 0.19 | 0.15 | | | |
| 60 | 0.33 | 0.25 | | | | 0.19 | 0.14 | | | |

Table SC.4. Of those who drank alcohol in the last week, proportion **heavy drinkers** (Males: 8+ units, Females: 6+ units on heaviest drinking day), by sex, birth cohort and age (3-year moving averages).

| | Males | | | | | Females | | | | |
|---|---|---|---|---|---|---|---|---|---|---|
| **Age** | **1940s** | **1950s** | **1960s** | **1970s** | **1980s** | **1940s** | **1950s** | **1960s** | **1970s** | **1980s** |
| 25 | | | | | 0.42 | | | | | 0.32 |
| 26 | | | | | 0.39 | | | | | 0.29 |
| 27 | | | | | 0.39 | | | | | 0.28 |
| 28 | | | | | 0.36 | | | | | 0.25 |
| 29 | | | | 0.39 | 0.33 | | | | 0.27 | 0.23 |
| 30 | | | | 0.37 | 0.30 | | | | 0.28 | 0.22 |
| 31 | | | | 0.37 | 0.29 | | | | 0.25 | 0.22 |
| 32 | | | | 0.34 | 0.30 | | | | 0.26 | 0.21 |
| 33 | | | | 0.34 | 0.28 | | | | 0.25 | 0.20 |
| 34 | | | | 0.34 | 0.27 | | | | 0.26 | 0.21 |
| 35 | | | | 0.32 | 0.25 | | | | 0.24 | 0.22 |
| 36 | | | | 0.31 | 0.25 | | | | 0.23 | 0.21 |
| 37 | | | | 0.30 | 0.23 | | | | 0.23 | 0.21 |
| 38 | | | | 0.31 | 0.23 | | | | 0.22 | 0.22 |
| 39 | | | 0.38 | 0.30 | | | | 0.26 | 0.21 | |
| 40 | | | 0.36 | 0.30 | | | | 0.25 | 0.20 | |
| 41 | | | 0.36 | 0.28 | | | | 0.26 | 0.21 | |
| 42 | | | 0.35 | 0.27 | | | | 0.25 | 0.20 | |
| 43 | | | 0.34 | 0.24 | | | | 0.26 | 0.20 | |
| 44 | | | 0.33 | 0.24 | | | | 0.24 | 0.19 | |
| 45 | | | 0.33 | 0.25 | | | | 0.24 | 0.21 | |
| 46 | | | 0.31 | 0.27 | | | | 0.23 | 0.22 | |
| 47 | | | 0.31 | 0.29 | | | | 0.22 | 0.20 | |
| 48 | | | 0.30 | 0.29 | | | | 0.21 | 0.22 | |
| 49 | | 0.32 | 0.30 | | | | 0.22 | 0.21 | | |
| 50 | | 0.35 | 0.30 | | | | 0.23 | 0.21 | | |
| 51 | | 0.33 | 0.29 | | | | 0.22 | 0.20 | | |
| 52 | | 0.30 | 0.29 | | | | 0.18 | 0.19 | | |
| 53 | | 0.29 | 0.26 | | | | 0.17 | 0.18 | | |
| 54 | | 0.27 | 0.28 | | | | 0.16 | 0.18 | | |
| 55 | | 0.27 | 0.27 | | | | 0.18 | 0.19 | | |
| 56 | | 0.25 | 0.30 | | | | 0.18 | 0.19 | | |
| 57 | | 0.26 | 0.28 | | | | 0.16 | 0.17 | | |
| 58 | | 0.24 | 0.26 | | | | 0.14 | 0.13 | | |
| 59 | 0.26 | 0.24 | | | | 0.13 | 0.15 | | | |
| 60 | 0.25 | 0.24 | | | | 0.14 | 0.15 | | | |

Table SC.5. Proportion **obese** (BMI 30kg/m$^2$ or higher) by sex, birth cohort and age (3-year moving averages).

| | Males | | | | | Females | | | | |
|---|---|---|---|---|---|---|---|---|---|---|
| **Age** | **1940s** | **1950s** | **1960s** | **1970s** | **1980s** | **1940s** | **1950s** | **1960s** | **1970s** | **1980s** |
| 25 | | | | 0.11 | 0.14 | | | | 0.14 | 0.20 |
| 26 | | | 0.09 | 0.12 | 0.17 | | | 0.11 | 0.14 | 0.20 |
| 27 | | | 0.11 | 0.14 | 0.18 | | | 0.12 | 0.14 | 0.19 |
| 28 | | | 0.11 | 0.16 | 0.18 | | | 0.13 | 0.15 | 0.20 |
| 29 | | | 0.12 | 0.17 | 0.19 | | | 0.14 | 0.18 | 0.23 |
| 30 | | | 0.13 | 0.19 | 0.19 | | | 0.14 | 0.20 | 0.25 |
| 31 | | | 0.13 | 0.20 | 0.19 | | | 0.15 | 0.21 | 0.25 |
| 32 | | | 0.15 | 0.20 | 0.20 | | | 0.16 | 0.22 | 0.25 |
| 33 | | | 0.16 | 0.21 | 0.21 | | | 0.18 | 0.22 | 0.25 |
| 34 | | | 0.17 | 0.21 | 0.24 | | | 0.18 | 0.23 | 0.25 |
| 35 | | | 0.19 | 0.22 | 0.25 | | | 0.21 | 0.23 | 0.26 |
| 36 | | 0.13 | 0.21 | 0.24 | 0.27 | | 0.16 | 0.21 | 0.24 | 0.27 |
| 37 | | 0.13 | 0.23 | 0.25 | 0.27 | | 0.16 | 0.22 | 0.24 | 0.29 |
| 38 | | 0.14 | 0.24 | 0.25 | 0.29 | | 0.18 | 0.21 | 0.25 | 0.35 |
| 39 | | 0.15 | 0.24 | 0.25 | | | 0.18 | 0.23 | 0.26 | |
| 40 | | 0.18 | 0.24 | 0.25 | | | 0.18 | 0.23 | 0.27 | |
| 41 | | 0.19 | 0.24 | 0.27 | | | 0.18 | 0.24 | 0.27 | |
| 42 | | 0.19 | 0.24 | 0.29 | | | 0.18 | 0.24 | 0.26 | |
| 43 | | 0.18 | 0.25 | 0.30 | | | 0.19 | 0.25 | 0.29 | |
| 44 | | 0.19 | 0.27 | 0.31 | | | 0.21 | 0.26 | 0.31 | |
| 45 | | 0.21 | 0.28 | 0.32 | | | 0.22 | 0.26 | 0.33 | |
| 46 | 0.17 | 0.23 | 0.30 | 0.34 | | 0.18 | 0.23 | 0.28 | 0.34 | |
| 47 | 0.21 | 0.25 | 0.31 | 0.37 | | 0.19 | 0.23 | 0.29 | 0.35 | |
| 48 | 0.22 | 0.26 | 0.31 | 0.35 | | 0.19 | 0.25 | 0.32 | 0.33 | |
| 49 | 0.21 | 0.28 | 0.31 | | | 0.20 | 0.26 | 0.32 | | |
| 50 | 0.20 | 0.29 | 0.31 | | | 0.22 | 0.27 | 0.32 | | |
| 51 | 0.22 | 0.30 | 0.33 | | | 0.23 | 0.28 | 0.32 | | |
| 52 | 0.22 | 0.32 | 0.34 | | | 0.25 | 0.29 | 0.32 | | |
| 53 | 0.23 | 0.32 | 0.35 | | | 0.25 | 0.30 | 0.32 | | |
| 54 | 0.23 | 0.34 | 0.34 | | | 0.26 | 0.29 | 0.34 | | |
| 55 | 0.25 | 0.33 | 0.33 | | | 0.26 | 0.30 | 0.33 | | |
| 56 | 0.26 | 0.35 | 0.36 | | | 0.27 | 0.30 | 0.36 | | |
| 57 | 0.27 | 0.35 | 0.37 | | | 0.28 | 0.32 | 0.34 | | |
| 58 | 0.28 | 0.36 | 0.36 | | | 0.30 | 0.33 | 0.34 | | |
| 59 | 0.30 | 0.34 | | | | 0.30 | 0.32 | | | |
| 60 | 0.29 | 0.34 | | | | 0.32 | 0.32 | | | |

Table SC.6. Mean BMI (kg/m$^2$), by sex, birth cohort and age (3-year moving averages).

| | Males | | | | | Females | | | | |
|---|---|---|---|---|---|---|---|---|---|---|
| **Age** | **1940s** | **1950s** | **1960s** | **1970s** | **1980s** | **1940s** | **1950s** | **1960s** | **1970s** | **1980s** |
| 25 | | | | 25.2 | 25.6 | | | | 24.8 | 25.8 |
| 26 | | | 25.1 | 25.6 | 26.0 | | | 24.6 | 25.0 | 25.9 |
| 27 | | | 25.3 | 25.9 | 26.2 | | | 24.8 | 25.2 | 26.0 |
| 28 | | | 25.4 | 26.2 | 26.4 | | | 24.9 | 25.4 | 26.2 |
| 29 | | | 25.6 | 26.3 | 26.5 | | | 25.0 | 25.7 | 26.5 |
| 30 | | | 25.8 | 26.5 | 26.5 | | | 25.1 | 26.1 | 26.8 |
| 31 | | | 26.0 | 26.7 | 26.5 | | | 25.2 | 26.3 | 26.8 |
| 32 | | | 26.2 | 26.8 | 26.8 | | | 25.4 | 26.5 | 26.9 |
| 33 | | | 26.4 | 26.9 | 27.0 | | | 25.7 | 26.6 | 27.0 |
| 34 | | | 26.5 | 27.1 | 27.4 | | | 25.9 | 26.7 | 27.0 |
| 35 | | | 26.7 | 27.3 | 27.3 | | | 26.2 | 26.6 | 27.0 |
| 36 | | 26.1 | 27.1 | 27.4 | 27.5 | | 25.6 | 26.3 | 26.8 | 27.2 |
| 37 | | 26.1 | 27.4 | 27.5 | 27.6 | | 25.6 | 26.5 | 26.9 | 27.6 |
| 38 | | 26.3 | 27.5 | 27.5 | 28.1 | | 25.7 | 26.5 | 27.2 | 28.4 |
| 39 | | 26.6 | 27.6 | 27.6 | | | 25.9 | 26.8 | 27.4 | |
| 40 | | 26.8 | 27.6 | 27.7 | | | 26.0 | 26.8 | 27.4 | |
| 41 | | 26.9 | 27.6 | 28.0 | | | 26.0 | 27.0 | 27.4 | |
| 42 | | 27.0 | 27.7 | 28.1 | | | 26.1 | 27.0 | 27.2 | |
| 43 | | 27.0 | 27.8 | 28.2 | | | 26.3 | 27.2 | 27.6 | |
| 44 | | 27.1 | 28.0 | 28.3 | | | 26.5 | 27.3 | 27.8 | |
| 45 | | 27.3 | 28.0 | 28.5 | | | 26.7 | 27.4 | 28.3 | |
| 46 | 26.6 | 27.5 | 28.3 | 28.9 | | 26.2 | 26.8 | 27.6 | 28.4 | |
| 47 | 27.0 | 27.6 | 28.5 | 29.0 | | 26.4 | 27.0 | 27.8 | 28.7 | |
| 48 | 27.2 | 27.8 | 28.6 | 28.9 | | 26.5 | 27.2 | 28.1 | 28.4 | |
| 49 | 27.2 | 27.9 | 28.6 | | | 26.7 | 27.5 | 28.3 | | |
| 50 | 27.2 | 28.1 | 28.5 | | | 26.9 | 27.6 | 28.4 | | |
| 51 | 27.4 | 28.2 | 28.6 | | | 27.0 | 27.7 | 28.4 | | |
| 52 | 27.5 | 28.4 | 28.7 | | | 27.2 | 27.9 | 28.4 | | |
| 53 | 27.6 | 28.5 | 28.8 | | | 27.3 | 27.9 | 28.3 | | |
| 54 | 27.6 | 28.6 | 28.8 | | | 27.4 | 27.9 | 28.3 | | |
| 55 | 27.7 | 28.6 | 28.6 | | | 27.5 | 28.1 | 28.3 | | |
| 56 | 27.8 | 28.7 | 28.9 | | | 27.6 | 28.2 | 28.4 | | |
| 57 | 27.9 | 28.8 | 29.0 | | | 27.8 | 28.3 | 28.6 | | |
| 58 | 28.1 | 28.9 | 29.2 | | | 27.9 | 28.3 | 28.3 | | |
| 59 | 28.2 | 28.8 | | | | 27.9 | 28.2 | | | |
| 60 | 28.2 | 28.8 | | | | 28.0 | 28.2 | | | |

Table SC.7. Proportion achieving 30+ minutes **moderate-intensity activity** on 5+ days/week (including activity whilst at work), by sex, birth cohort and age (3-year moving averages).

| | Males | | | | | Females | | | | |
|---|---|---|---|---|---|---|---|---|---|---|
| **Age** | **1940s** | **1950s** | **1960s** | **1970s** | **1980s** | **1940s** | **1950s** | **1960s** | **1970s** | **1980s** |
| 25 | | | | 0.48 | 0.56 | | | | 0.31 | 0.38 |
| 26 | | | | 0.47 | 0.56 | | | | 0.32 | 0.37 |
| 27 | | | | 0.46 | 0.58 | | | | 0.32 | 0.40 |
| 28 | | | | 0.48 | 0.57 | | | | 0.33 | 0.41 |
| 29 | | | 0.44 | 0.49 | 0.59 | | | 0.30 | 0.34 | 0.40 |
| 30 | | | 0.46 | 0.50 | 0.60 | | | 0.28 | 0.34 | 0.37 |
| 31 | | | 0.46 | 0.48 | 0.58 | | | 0.27 | 0.34 | 0.38 |
| 32 | | | 0.45 | 0.48 | | | | 0.29 | 0.34 | |
| 33 | | | 0.44 | 0.49 | | | | 0.29 | 0.34 | |
| 34 | | | 0.41 | 0.47 | | | | 0.31 | 0.33 | |
| 35 | | | 0.43 | 0.46 | | | | 0.31 | 0.33 | |
| 36 | | | 0.43 | 0.48 | | | | 0.32 | 0.36 | |
| 37 | | | 0.45 | 0.50 | | | | 0.31 | 0.37 | |
| 38 | | | 0.43 | 0.53 | | | | 0.31 | 0.42 | |
| 39 | | 0.41 | 0.44 | 0.49 | | | 0.28 | 0.33 | 0.38 | |
| 40 | | 0.43 | 0.45 | 0.48 | | | 0.29 | 0.35 | 0.37 | |
| 41 | | 0.41 | 0.45 | 0.43 | | | 0.31 | 0.36 | 0.36 | |
| 42 | | 0.40 | 0.44 | | | | 0.32 | 0.36 | | |
| 43 | | 0.39 | 0.43 | | | | 0.34 | 0.35 | | |
| 44 | | 0.39 | 0.44 | | | | 0.32 | 0.35 | | |
| 45 | | 0.37 | 0.45 | | | | 0.31 | 0.37 | | |
| 46 | | 0.37 | 0.46 | | | | 0.31 | 0.38 | | |
| 47 | | 0.36 | 0.47 | | | | 0.32 | 0.38 | | |
| 48 | | 0.38 | 0.43 | | | | 0.33 | 0.36 | | |
| 49 | 0.33 | 0.39 | 0.42 | | | 0.31 | 0.32 | 0.39 | | |
| 50 | 0.36 | 0.39 | 0.40 | | | 0.27 | 0.31 | 0.38 | | |
| 51 | 0.36 | 0.38 | 0.43 | | | 0.28 | 0.30 | 0.34 | | |
| 52 | 0.36 | 0.35 | | | | 0.29 | 0.32 | | | |
| 53 | 0.37 | 0.35 | | | | 0.29 | 0.31 | | | |
| 54 | 0.37 | 0.36 | | | | 0.28 | 0.33 | | | |
| 55 | 0.36 | 0.37 | | | | 0.26 | 0.30 | | | |
| 56 | 0.34 | 0.36 | | | | 0.25 | 0.31 | | | |
| 57 | 0.34 | 0.34 | | | | 0.23 | 0.30 | | | |
| 58 | 0.34 | 0.32 | | | | 0.25 | 0.34 | | | |
| 59 | 0.34 | 0.33 | | | | 0.25 | 0.32 | | | |
| 60 | 0.33 | 0.31 | | | | 0.25 | 0.34 | | | |

Table SC.8. Proportion consuming 5+ **fruit and vegetables** per day, by sex, birth cohort and age (3-year moving averages).

| | Males | | | | | Females | | | | |
|---|---|---|---|---|---|---|---|---|---|---|
| **Age** | **1940s** | **1950s** | **1960s** | **1970s** | **1980s** | **1940s** | **1950s** | **1960s** | **1970s** | **1980s** |
| 25 | | | | 0.16 | 0.19 | | | | 0.22 | 0.24 |
| 26 | | | | 0.18 | 0.20 | | | | 0.24 | 0.24 |
| 27 | | | | 0.19 | 0.19 | | | | 0.25 | 0.25 |
| 28 | | | | 0.21 | 0.22 | | | | 0.26 | 0.25 |
| 29 | | | | 0.23 | 0.23 | | | | 0.27 | 0.26 |
| 30 | | | | 0.23 | 0.24 | | | | 0.27 | 0.27 |
| 31 | | | | 0.24 | 0.24 | | | | 0.28 | 0.29 |
| 32 | | | | 0.25 | 0.26 | | | | 0.28 | 0.29 |
| 33 | | | 0.21 | 0.26 | | | | 0.26 | 0.28 | |
| 34 | | | 0.23 | 0.26 | | | | 0.25 | 0.28 | |
| 35 | | | 0.24 | 0.26 | | | | 0.26 | 0.29 | |
| 36 | | | 0.23 | 0.26 | | | | 0.28 | 0.30 | |
| 37 | | | 0.24 | 0.27 | | | | 0.28 | 0.30 | |
| 38 | | | 0.24 | 0.25 | | | | 0.29 | 0.32 | |
| 39 | | | 0.24 | 0.26 | | | | 0.29 | 0.32 | |
| 40 | | | 0.24 | 0.25 | | | | 0.29 | 0.30 | |
| 41 | | | 0.24 | 0.27 | | | | 0.29 | 0.30 | |
| 42 | | | 0.25 | 0.25 | | | | 0.29 | 0.30 | |
| 43 | | 0.20 | 0.26 | | | | 0.28 | 0.28 | | |
| 44 | | 0.21 | 0.27 | | | | 0.27 | 0.29 | | |
| 45 | | 0.23 | 0.27 | | | | 0.27 | 0.29 | | |
| 46 | | 0.23 | 0.26 | | | | 0.28 | 0.30 | | |
| 47 | | 0.24 | 0.25 | | | | 0.30 | 0.29 | | |
| 48 | | 0.25 | 0.23 | | | | 0.33 | 0.28 | | |
| 49 | | 0.26 | 0.23 | | | | 0.33 | 0.29 | | |
| 50 | | 0.27 | 0.22 | | | | 0.34 | 0.28 | | |
| 51 | | 0.27 | 0.21 | | | | 0.35 | 0.30 | | |
| 52 | | 0.28 | 0.23 | | | | 0.36 | 0.30 | | |
| 53 | 0.27 | 0.28 | | | | 0.34 | 0.35 | | | |
| 54 | 0.28 | 0.29 | | | | 0.36 | 0.34 | | | |
| 55 | 0.27 | 0.29 | | | | 0.37 | 0.33 | | | |
| 56 | 0.27 | 0.29 | | | | 0.36 | 0.33 | | | |
| 57 | 0.27 | 0.27 | | | | 0.36 | 0.34 | | | |
| 58 | 0.30 | 0.26 | | | | 0.36 | 0.33 | | | |
| 59 | 0.31 | 0.25 | | | | 0.36 | 0.32 | | | |
| 60 | 0.31 | 0.23 | | | | 0.36 | 0.34 | | | |

#### D: Tables of predicted prevalence and prevalence ratios by Index of Multiple Deprivation (IMD) quintiles at age 40 (absolute and relative inequalities)

Table SD1. Current smoking : absolute and relative inequalities at age 40, by sex and cohort

| | 1940s | 1950s | 1960s | 1970s | 1980s | Change (%) |
|---|---|---|---|---|---|---|
| ***Males*** | | | | | | |
| Most deprived (Q1) | 0.59 | 0.53 | 0.42 | 0.35 | 0.31 | *-48%* |
| Q2 | 0.46 | 0.38 | 0.33 | 0.30 | 0.23 | *-50%* |
| Q3 | 0.36 | 0.31 | 0.27 | 0.25 | 0.23 | *-36%* |
| Q4 | 0.28 | 0.25 | 0.23 | 0.21 | 0.19 | *-33%* |
| Least deprived (Q5) | 0.23 | 0.19 | 0.19 | 0.18 | 0.15 | *-32%* |
| **Absolute inequality:** | | | | | | |
| **PP[1] difference (Q1-Q5)** | 0.36 | 0.34 | 0.23 | 0.17 | 0.15 | |
| **Relative inequality: Prev Ratio (PR) (Q1/Q5)** | 2.62 | 2.79 | 2.19 | 1.95 | 2.00 | |
| 95% CI of PR | *2.34,2.92* | *2.57,3.04* | *2.04,2.35* | *1.78,2.14* | *1.70,2.34* | |
| PR(C[2])/PR (1940) | 1 | 1.07 | 0.84 | 0.75 | 0.76 | |
| 95% CI | | *0.93,1.23* | *0.74,0.95* | *0.65,0.86* | *0.63,0.92* | |
| P | | 0.355 | 0.008 | <0.001 | 0.006 | |
| P[3] (for 4 terms) | <0.001 | | | | | |
| ***Females*** | | | | | | |
| Most deprived (Q1) | 0.56 | 0.51 | 0.39 | 0.29 | 0.24 | *-57%* |
| Q2 | 0.44 | 0.38 | 0.31 | 0.26 | 0.19 | *-58%* |
| Q3 | 0.35 | 0.30 | 0.25 | 0.20 | 0.16 | *-54%* |
| Q4 | 0.28 | 0.23 | 0.19 | 0.17 | 0.14 | *-51%* |
| Least deprived (Q5) | 0.24 | 0.18 | 0.15 | 0.12 | 0.11 | *-55%* |
| **Absolute inequality:** | | | | | | |
| **PP[1] difference (Q1-Q5)** | 0.32 | 0.33 | 0.24 | 0.17 | 0.14 | |
| **Relative inequality: Prev Ratio (PR) (Q1/Q5)** | 2.35 | 2.81 | 2.58 | 2.38 | 2.27 | |
| 95% CI of PR | *2.12,2.60* | *2.60,3.05* | *2.40,2.77* | *2.16,2.63* | *1.92,2.68* | |
| PR(C[2])/PR (1940) | 1 | 1.20 | 1.10 | 1.02 | 0.97 | |
| 95% CI | | *1.05,1.37* | *0.97,1.24* | *0.88,1.17* | *0.79,1.17* | |
| P | | 0.005 | 0.142 | 0.831 | 0.728 | |
| P[3] (for 4 terms) | 0.017 | | | | | |

Notes: Predicted prevalence in table expressed as a proportion. Log-binomial regression model : age + cohort + IMD + (cohort x IMD). IMD fitted as 4 indicator variables.

[1]Percentage Point (PP) difference

[2] PR for <cohort>/PR for ref cohort (1940=1). If the estimated PR value and its 95% CI does not include 1, this indicates a significant difference in the estimated PR for that specific cohort versus the 1940s cohort.

[3] P shown for interaction term testing change in relative inequality (4 PRs specifically comparing the least and most deprived quintiles) between cohorts (Joint significance test; 1940s cohort as reference).

Table SD2. Heavy smoking : absolute and relative inequalities at age 40, by sex and cohort

| | 1940s | 1950s | 1960s | 1970s | 1980s | Change (%) |
|---|---|---|---|---|---|---|
| ***Males*** | | | | | | |
| Most deprived (Q1) | 0.59 | 0.53 | 0.40 | 0.28 | 0.15 | *-74%* |
| Q2 | 0.54 | 0.48 | 0.35 | 0.24 | 0.19 | *-64%* |
| Q3 | 0.54 | 0.46 | 0.33 | 0.22 | 0.14 | *-75%* |
| Q4 | 0.51 | 0.44 | 0.31 | 0.21 | 0.15 | *-70%* |
| Least deprived (Q5) | 0.48 | 0.42 | 0.29 | 0.21 | 0.11 | *-77%* |
| **Absolute inequality:** | | | | | | |
| **PP[1] difference (Q1-Q5)** | 0.11 | 0.11 | 0.11 | 0.07 | 0.04 | |
| **Relative inequality: Prev** | | | | | | |
| **Ratio (PR) (Q1/Q5)** | 1.23 | 1.26 | 1.37 | 1.31 | 1.34 | |
| 95% CI of PR | *1.08,1.39* | *1.12,1.41* | *1.21,1.56* | *1.07,1.61* | *0.71,2.54* | |
| PR(C[2])/PR (1940) | 1 | 1.02 | 1.12 | 1.07 | 1.09 | |
| 95% CI | | *0.86,1.21* | *0.93,1.34* | *0.8,1.33* | *0.57,2.09* | |
| P | | 0.79 | 0.23 | 0.59 | 0.79 | |
| P[3] (for 4 terms) | 0.792 | | | | | |
| ***Females*** | | | | | | |
| Most deprived (Q1) | 0.48 | 0.45 | 0.34 | 0.24 | 0.13 | *-73%* |
| Q2 | 0.44 | 0.40 | 0.28 | 0.19 | 0.11 | *-76%* |
| Q3 | 0.40 | 0.32 | 0.25 | 0.17 | 0.12 | *-70%* |
| Q4 | 0.33 | 0.32 | 0.22 | 0.13 | 0.06 | *-82%* |
| Least deprived (Q5) | 0.36 | 0.26 | 0.17 | 0.11 | 0.05 | *-86%* |
| **Absolute inequality:** | | | | | | |
| **PP[1] difference (Q1-Q5)** | 0.12 | 0.19 | 0.17 | 0.13 | 0.08 | |
| **Relative inequality: Prev** | | | | | | |
| **Ratio (PR) (Q1/Q5)** | 1.34 | 1.72 | 2.01 | 2.16 | 2.55 | |
| 95% CI of PR | *1.15,1.57* | *1.48,1.99* | *1.7,2.38* | *1.62,2.9* | *1.05,6.2* | |
| PR(C[2])/PR (1940) | 1 | 1.28 | 1.49 | 1.61 | 1.89 | |
| 95% CI | | *1.03,1.58* | *1.19,1.88* | *1.16,2.24* | *0.77,4.68* | |
| P | | 0.025 | 0.001 | 0.005 | 0.166 | |
| P[3] (for 4 terms) | 0.003 | | | | | |

Notes: Predicted prevalence in table expressed as a proportion. Log-binomial regression model : age + cohort + IMD + (cohort x IMD). IMD fitted as 4 indicator variables.

[1]Percentage Point (PP) difference

[2] PR for <cohort>/PR for ref cohort (1940=1). If the estimated PR value and its 95% CI does not include 1, this indicates a significant difference in the estimated PR for that specific cohort versus the 1940s cohort.

[3] P shown for interaction term testing change in relative inequality (4 PRs specifically comparing the least and most deprived quintiles) between cohorts (Joint significance test; 1940s cohort as reference).

Table SD3. Alcohol 5+ days/week : absolute and relative inequalities at age 40, by sex and cohort

| | 1940s | 1950s | 1960s | 1970s | 1980s | Change (%) |
|---|---|---|---|---|---|---|
| ***Males*** | | | | | | |
| Most deprived (Q1) | 0.28 | 0.21 | 0.17 | 0.10 | 0.07 | *-74%* |
| Q2 | 0.29 | 0.24 | 0.17 | 0.15 | 0.08 | *-71%* |
| Q3 | 0.35 | 0.26 | 0.19 | 0.14 | 0.10 | *-72%* |
| Q4 | 0.34 | 0.30 | 0.21 | 0.15 | 0.07 | *-78%* |
| Least deprived (Q5) | 0.38 | 0.31 | 0.24 | 0.15 | 0.08 | *-79%* |
| **Absolute inequality:** | | | | | | |
| **PP[1] difference (Q1-Q5)** | -0.10 | -0.10 | -0.07 | -0.05 | -0.01 | |
| **Relative inequality: Prev Ratio (PR) (Q1/Q5)** | 0.75 | 0.68 | 0.72 | 0.70 | 0.89 | |
| 95% CI of PR | *0.68,0.83* | *0.63,0.75* | *0.65,0.78* | *0.61,0.81* | *0.67,1.20* | |
| PR(C[2])/PR (1940) | 1 | 0.92 | 0.96 | 0.94 | 1.20 | |
| 95% CI | | *0.80,1.05* | *0.84,1.10* | *0.79,1.12* | *0.88,1.63* | |
| P | | 0.203 | 0.536 | 0.462 | 0.255 | |
| P[3] (for 4 terms) | 0.413 | | | | | |
| ***Females*** | | | | | | |
| Most deprived (Q1) | 0.11 | 0.11 | 0.08 | 0.05 | 0.03 | *-78%* |
| Q2 | 0.15 | 0.13 | 0.10 | 0.07 | 0.02 | *-84%* |
| Q3 | 0.19 | 0.16 | 0.12 | 0.08 | 0.04 | *-79%* |
| Q4 | 0.21 | 0.20 | 0.14 | 0.09 | 0.04 | *-81%* |
| Least deprived (Q5) | 0.23 | 0.20 | 0.15 | 0.09 | 0.04 | *-81%* |
| **Absolute inequality:** | | | | | | |
| **PP[1] difference (Q1-Q5)** | -0.12 | -0.09 | -0.07 | -0.04 | -0.02 | |
| **Relative inequality: Prev Ratio (PR) (Q1/Q5)** | 0.49 | 0.53 | 0.55 | 0.56 | 0.57 | |
| 95% CI of PR | *0.42,0.57* | *0.47,0.60* | *0.49,0.62* | *0.46,0.67* | *0.39,0.84* | |
| PR(C[2])/PR (1940) | 1 | 1.08 | 1.12 | 1.14 | 1.17 | |
| 95% CI | | *0.89,1.31* | *0.92,1.35* | *0.90,1.44* | *0.77,1.75* | |
| P | | 0.420 | 0.257 | 0.290 | 0.464 | |
| P[3] (for 4 terms) | 0.784 | | | | | |

Notes: Predicted prevalence in table expressed as a proportion. Log-binomial regression model : age + cohort + IMD + (cohort x IMD). IMD fitted as 4 indicator variables.

[1]Percentage Point (PP) difference

[2] PR for <cohort>/PR for ref cohort (1940=1). If the estimated PR value and its 95% CI does not include 1, this indicates a significant difference in the estimated PR for that specific cohort versus the 1940s cohort.

[3] P shown for interaction term testing change in relative inequality (4 PRs specifically comparing the least and most deprived quintiles) between cohorts (Joint significance test; 1940s cohort as reference).

Table SD4. Heavy drinking : absolute and relative inequalities at age 40, by sex and cohort

| | 1940s | 1950s | 1960s | 1970s | 1980s | Change (%) |
|---|---|---|---|---|---|---|
| ***Males*** | | | | | | |
| Most deprived (Q1) | 0.43 | 0.52 | 0.48 | 0.36 | 0.32 | *-27%* |
| Q2 | 0.39 | 0.50 | 0.46 | 0.37 | 0.30 | *-23%* |
| Q3 | 0.54 | 0.44 | 0.46 | 0.35 | 0.29 | *-46%* |
| Q4 | 0.42 | 0.44 | 0.40 | 0.35 | 0.31 | *-25%* |
| Least deprived (Q5) | 0.39 | 0.40 | 0.39 | 0.31 | 0.31 | *-22%* |
| **Absolute inequality:** | | | | | | |
| **PP[1] difference (Q1-Q5)** | 0.04 | 0.12 | 0.09 | 0.05 | 0.01 | |
| **Relative inequality: Prev Ratio (PR) (Q1/Q5)** | 1.10 | 1.31 | 1.24 | 1.16 | 1.03 | |
| 95% CI of PR | *0.69,1.75* | *1.13,1.52* | *1.11,1.39* | *1.03,1.31* | *0.90,1.19* | |
| PR(C[2])/PR (1940) | 1 | 1.19 | 1.13 | 1.06 | 0.94 | |
| 95% CI | | *0.73,1.95* | *0.70,1.83* | *0.65,1.71* | *0.58,1.53* | |
| P | | 0.486 | 0.621 | 0.822 | 0.812 | |
| P[3] (for 4 terms) | 0.179 | | | | | |
| ***Females*** | | | | | | |
| Most deprived (Q1) | 0.36 | 0.38 | 0.39 | 0.36 | 0.33 | *-8%* |
| Q2 | 0.40 | 0.30 | 0.34 | 0.28 | 0.28 | *-31%* |
| Q3 | 0.18 | 0.27 | 0.34 | 0.28 | 0.25 | *33%* |
| Q4 | 0.24 | 0.30 | 0.31 | 0.29 | 0.26 | *8%* |
| Least deprived (Q5) | 0.32 | 0.28 | 0.28 | 0.23 | 0.25 | *-21%* |
| **Absolute inequality:** | | | | | | |
| **PP[1] difference (Q1-Q5)** | 0.04 | 0.09 | 0.12 | 0.13 | 0.08 | |
| **Relative inequality: Prev Ratio (PR) (Q1/Q5)** | 1.11 | 1.34 | 1.42 | 1.54 | 1.30 | |
| 95% CI of PR | *0.59,2.10* | *1.10,1.62* | *1.25,1.61* | *1.35,1.75* | *1.13,1.50* | |
| PR(C[2])/PR (1940) | 1 | 1.20 | 1.27 | 1.38 | 1.17 | |
| 95% CI | | *0.62,2.33* | *0.67,2.43* | *0.72,2.63* | *0.61,2.23* | |
| P | | 0.591 | 0.465 | 0.327 | 0.639 | |
| P[3] (for 4 terms) | 0.432 | | | | | |

Notes: Predicted prevalence in table expressed as a proportion. Log-binomial regression model : age + cohort + IMD + (cohort x IMD). IMD fitted as 4 indicator variables.

[1] Percentage Point (PP) difference

[2] PR for <cohort>/PR for ref cohort (1940=1). If the estimated PR value and its 95% CI does not include 1, this indicates a significant difference in the estimated PR for that specific cohort versus the 1940s cohort.

[3] P shown for interaction term testing change in relative inequality (4 PRs specifically comparing the least and most deprived quintiles) between cohorts (Joint significance test; 1940s cohort as reference).

Table SD5. Obesity (BMI 30kg/m$^2$+): absolute and relative inequalities at age 40, by sex and cohort

| | 1940s | 1950s | 1960s | 1970s | 1980s | Change (%) |
|---|---|---|---|---|---|---|
| ***Males*** | | | | | | |
| Most deprived (Q1) | 0.15 | 0.19 | 0.23 | 0.27 | 0.31 | *99%* |
| Q2 | 0.15 | 0.20 | 0.23 | 0.28 | 0.28 | *84%* |
| Q3 | 0.14 | 0.18 | 0.23 | 0.25 | 0.30 | *117%* |
| Q4 | 0.13 | 0.17 | 0.22 | 0.26 | 0.26 | *98%* |
| Least deprived (Q5) | 0.12 | 0.16 | 0.20 | 0.23 | 0.27 | *127%* |
| **Absolute inequality: PP[1] difference (Q1-Q5)** | 0.03 | 0.04 | 0.03 | 0.04 | 0.04 | |
| **Relative inequality: Prev Ratio (PR) (Q5/Q1)** | 1.29 | 1.24 | 1.17 | 1.17 | 1.13 | |
| 95% CI of PR | *1.15,1.45* | *1.14,1.35* | *1.08,1.27* | *1.04,1.30* | *0.94,1.36* | |
| PR(C[2])/PR (1940) | 1 | 0.96 | 0.91 | 0.91 | 0.88 | |
| 95% CI | | *0.83,1.11* | *0.79,1.05* | *0.77,1.06* | *0.70,1.09* | |
| P | | 0.601 | 0.194 | 0.225 | 0.235 | |
| P[3] (for 4 terms) | 0.573 | | | | | |
| ***Females*** | | | | | | |
| Most deprived (Q1) | 0.20 | 0.24 | 0.29 | 0.35 | 0.44 | *120%* |
| Q2 | 0.18 | 0.20 | 0.26 | 0.30 | 0.33 | *81%* |
| Q3 | 0.16 | 0.18 | 0.23 | 0.25 | 0.31 | *91%* |
| Q4 | 0.14 | 0.17 | 0.19 | 0.21 | 0.27 | *90%* |
| Least deprived (Q5) | 0.13 | 0.13 | 0.16 | 0.19 | 0.24 | *85%* |
| **Absolute inequality: PP[1] difference (Q1-Q5)** | 0.07 | 0.10 | 0.14 | 0.16 | 0.20 | |
| **Relative inequality: Prev Ratio (PR) (Q5/Q1)** | 1.55 | 1.75 | 1.87 | 1.83 | 1.85 | |
| 95% CI of PR | *1.40,1.72* | *1.62,1.90* | *1.73,2.01* | *1.66,2.03* | *1.58,2.16* | |
| PR(C[2])/PR (1940) | 1 | 1.13 | 1.20 | 1.18 | 1.19 | |
| 95% CI | | *0.99,1.29* | *1.06,1.37* | *1.03,1.36* | *0.99,1.44* | |
| P | | 0.064 | 0.004 | 0.021 | 0.069 | |
| P[3] (for 4 terms) | 0.055 | | | | | |

Notes: Predicted prevalence in table expressed as a proportion. Log-binomial regression model : age + cohort + IMD + (cohort x IMD). IMD fitted as 4 indicator variables.

[1]Percentage Point (PP) difference

[2] PR for <cohort>/PR for ref cohort (1940=1). If the estimated PR value and its 95% CI does not include 1, this indicates a significant difference in the estimated PR for that specific cohort versus the 1940s cohort.

[3] P shown for interaction term testing change in relative inequality (4 PRs specifically comparing the least and most deprived quintiles) between cohorts (Joint significance test; 1940s cohort as reference).

Table SD6. Mean BMI: absolute inequalities at age 40, by sex and cohort

| | 1940s | 1950s | 1960s | 1970s | 1980s | Change (%) |
|---|---|---|---|---|---|---|
| ***Males*** | | | | | | |
| Most deprived (Q1) | 25.8 | 26.6 | 27.3 | 27.7 | 27.9 | *8%* |
| Q2 | 26.0 | 26.9 | 27.4 | 27.9 | 27.8 | *7%* |
| Q3 | 25.8 | 26.7 | 27.5 | 27.7 | 27.9 | *8%* |
| Q4 | 25.7 | 26.6 | 27.4 | 27.9 | 27.8 | *8%* |
| Least deprived (Q5) | 25.5 | 26.4 | 27.2 | 27.6 | 27.9 | *9%* |
| **Absolute inequality (gap):** | | | | | | |
| **Mean[1] difference (Q1-Q5)** | 0.31 | 0.19 | 0.02 | 0.11 | -0.02 | |
| 95% CI of Gap | *0.03,0.60* | *-0.03,0.42* | *-0.18,0.22* | *-0.14,0.36* | *-0.42,0.38* | |
| Gap (C[2])- Gap (1940) | 0 | -0.12 | -0.29 | -0.20 | -0.34 | |
| Interactions 95% CI | | -0.49,0.24 | -0.64,0.06 | -0.59,0.18 | -0.83,0.16 | |
| P | | 0.510 | 0.103 | 0.299 | 0.182 | |
| P[3] (for 4 terms) | 0.474 | | | | | |
| ***Females*** | | | | | | |
| Most deprived (Q1) | 26.3 | 27.1 | 27.9 | 28.4 | 29.2 | *11%* |
| Q2 | 25.9 | 26.5 | 27.3 | 27.8 | 28.0 | *8%* |
| Q3 | 25.5 | 25.9 | 26.8 | 27.2 | 27.8 | *9%* |
| Q4 | 25.0 | 25.6 | 26.2 | 26.6 | 27.4 | *10%* |
| Least deprived (Q5) | 24.7 | 25.1 | 25.7 | 26.4 | 26.9 | *9%* |
| **Absolute inequality (gap):** | | | | | | |
| **Mean[1] difference (Q1-Q5)** | 1.59 | 1.99 | 2.11 | 1.94 | 2.26 | |
| 95% CI of Gap | *1.25,1.94* | *1.72,2.25* | *1.88,2.33* | *1.65,2.22* | *1.82,2.71* | |
| Gap (C[2])- Gap (1940) | 0 | 0.39 | 0.51 | 0.34 | 0.67 | |
| Interactions 95% CI | | *-0.04,0.83* | *0.10,0.92* | *-0.10,0.79* | *0.10,1.23* | |
| P | | 0.079 | 0.015 | 0.132 | 0.020 | |
| P[3] (for 4 terms) | 0.106 | | | | | |

Notes: Predicted mean BMI at age 40. Linear regression model : age + cohort + IMD + (cohort x IMD). IMD fitted as 4 indicator variables.

[1]Mean difference

[2] Mean for <cohort> minus mean for ref cohort (1940=1). If the estimated mean difference and its 95% CI does not include 0, this indicates a significant difference in mean BMI between the specific cohort versus the 1940s cohort.

[3] P shown for interaction term testing change in absolute inequality (4 terms comparing the difference in mean BMI between the least and deprived quintiles) between cohorts (Joint significance test; 1940s cohort as reference).

Table SD7. Achieving sufficient levels of moderate-intensity physical activity: absolute and relative inequalities at age 40, by sex and cohort

| | 1940s | 1950s | 1960s | 1970s | 1980s | Change (%) |
|---|---|---|---|---|---|---|
| ***Males*** | | | | | | |
| Most deprived (Q1) | 0.32 | 0.36 | 0.41 | 0.47 | 0.49 | *56%* |
| Q2 | 0.40 | 0.41 | 0.46 | 0.49 | 0.59 | *46%* |
| Q3 | 0.38 | 0.40 | 0.47 | 0.49 | 0.57 | *49%* |
| Q4 | 0.36 | 0.40 | 0.45 | 0.43 | 0.60 | *68%* |
| Least deprived (Q5) | 0.34 | 0.35 | 0.40 | 0.42 | 0.48 | *40%* |
| **Absolute inequality: PP[1] difference (Q1-Q5)** | -0.03 | 0.00 | 0.01 | 0.05 | 0.01 | |
| **Relative inequality: Prev Ratio (PR) (Q5/Q1)** | 0.92 | 1.01 | 1.03 | 1.12 | 1.02 | |
| 95% CI of PR | *0.79,1.08* | *0.90,1.14* | *0.94,1.13* | *1.00,1.25* | *0.84,1.26* | |
| PR(C[2])/PR (1940) | 1 | 1.10 | 1.12 | 1.21 | 1.11 | |
| 95% CI | | *0.90,1.34* | *0.93,1.34* | *1.00,1.47* | *0.86,1.44* | |
| P | | 0.358 | 0.247 | 0.049 | 0.423 | |
| P[3] (for 4 terms) | 0.383 | | | | | |
| ***Females*** | | | | | | |
| Most deprived (Q1) | 0.20 | 0.27 | 0.31 | 0.34 | 0.42 | *111%* |
| Q2 | 0.24 | 0.30 | 0.35 | 0.39 | 0.42 | *77%* |
| Q3 | 0.23 | 0.29 | 0.34 | 0.38 | 0.42 | *84%* |
| Q4 | 0.24 | 0.31 | 0.32 | 0.37 | 0.49 | *102%* |
| Least deprived (Q5) | 0.23 | 0.29 | 0.35 | 0.33 | 0.47 | *108%* |
| **Absolute inequality: PP[1] difference (Q1-Q5)** | -0.03 | -0.02 | -0.04 | 0.02 | -0.05 | |
| **Relative inequality: Prev Ratio (PR) (Q5/Q1)** | 0.89 | 0.94 | 0.88 | 1.05 | 0.90 | |
| 95% CI of PR | *0.74,1.06* | *0.84,1.06* | *0.80,0.98* | *0.92,1.19* | *0.73,1.11* | |
| PR(C[2])/PR (1940) | 1 | 1.06 | 1.00 | 1.18 | 1.02 | |
| 95% CI | | *0.86,1.32* | *0.81,1.22* | *0.94,1.47* | *0.77,1.34* | |
| P | | 0.588 | 0.967 | 0.146 | 0.908 | |
| P[3] (for 4 terms) | 0.329 | | | | | |

Notes: Predicted prevalence in table expressed as a proportion. Log-binomial regression model : age + cohort + IMD + (cohort x IMD). IMD fitted as 4 indicator variables.

[1]Percentage Point (PP) difference

[2] PR for <cohort>/PR for ref cohort (1940=1). If the estimated PR value and its 95% CI does not include 1, this indicates a significant difference in the estimated PR for that specific cohort versus the 1940s cohort.

[3] P shown for interaction term testing change in relative inequality (4 PRs specifically comparing the least and most deprived quintiles) between cohorts (Joint significance test; 1940s cohort as reference).

Table SD8. 5+ fruit and vegetables/day: absolute and relative inequalities at age 40, by sex and cohort

| | 1940s | 1950s | 1960s | 1970s | 1980s | Change (%) |
|---|---|---|---|---|---|---|
| ***Males*** | | | | | | |
| Most deprived (Q1) | 0.15 | 0.17 | 0.21 | 0.22 | 0.19 | *28%* |
| Q2 | 0.19 | 0.20 | 0.24 | 0.26 | 0.28 | *44%* |
| Q3 | 0.21 | 0.21 | 0.23 | 0.26 | 0.27 | *25%* |
| Q4 | 0.27 | 0.24 | 0.24 | 0.26 | 0.29 | *7%* |
| Least deprived (Q5) | 0.28 | 0.26 | 0.26 | 0.30 | 0.27 | *-4%* |
| **Absolute inequality:** | | | | | | |
| **PP[1] difference (Q1-Q5)** | -0.13 | -0.08 | -0.05 | -0.08 | -0.08 | |
| **Relative inequality: Prev Ratio (PR) (Q5/Q1)** | 0.52 | 0.67 | 0.81 | 0.74 | 0.69 | |
| 95% CI of PR | *0.43,0.63* | *0.59,0.77* | *0.71,0.91* | *0.65,0.85* | *0.52,0.92* | |
| PR(C[2])/PR (1940) | 1 | 1.29 | 1.55 | 1.43 | 1.33 | |
| 95% CI | | *1.02,1.63* | *1.23,1.95* | *1.12,1.81* | *0.94,1.88* | |
| P | | 0.032 | <0.001 | 0.003 | 0.105 | |
| P[3] (for 4 terms | 0.005 | | | | | |
| ***Females*** | | | | | | |
| Most deprived (Q1) | 0.20 | 0.20 | 0.22 | 0.24 | 0.23 | *15%* |
| Q2 | 0.25 | 0.26 | 0.26 | 0.27 | 0.30 | *21%* |
| Q3 | 0.32 | 0.29 | 0.27 | 0.31 | 0.29 | *-9%* |
| Q4 | 0.35 | 0.32 | 0.30 | 0.34 | 0.29 | *-17%* |
| Least deprived (Q5) | 0.36 | 0.35 | 0.32 | 0.35 | 0.30 | *-17%* |
| **Absolute inequality:** | | | | | | |
| **PP[1] difference (Q1-Q5)** | -0.16 | -0.16 | -0.11 | -0.11 | -0.07 | |
| **Relative inequality: Prev Ratio (PR) (Q5/Q1)** | 0.56 | 0.56 | 0.67 | 0.69 | 0.78 | |
| 95% CI of PR | *0.48,0.65* | *0.50,0.63* | *0.60,0.74* | *0.62,0.76* | *0.63,0.97* | |
| PR(C[2])/PR (1940) | 1 | 1.00 | 1.20 | 1.23 | 1.39 | |
| 95% CI | | *0.83,1.21* | *1.00,1.44* | *1.02,1.48* | *1.07,1.82* | |
| P | | 0.980 | 0.053 | 0.031 | 0.014 | |
| P[3] (for 4 terms) | 0.008 | | | | | |

Notes: Predicted prevalence in table expressed as a proportion. Log-binomial regression model : age + cohort + IMD + (cohort x IMD). IMD fitted as 4 indicator variables.

[1]Percentage Point (PP) difference

[2] PR for <cohort>/PR for ref cohort (1940=1). If the estimated PR value and its 95% CI does not include 1, this indicates a significant difference in the estimated PR for that specific cohort versus the 1940s cohort.

[3] P shown for interaction term testing change in relative inequality (4 PRs specifically comparing the least and most deprived quintiles) between cohorts (Joint significance test; 1940s cohort as reference).
